# Inferring temporal trends of multiple pathogens, variants, subtypes or serotypes from routine surveillance data

**DOI:** 10.1101/2024.11.03.24316681

**Authors:** Oliver Eales, Saras M. Windecker, James M. McCaw, Freya M. Shearer

## Abstract

Estimating the temporal trends in infectious disease activity is crucial for monitoring disease spread and the impact of interventions. Surveillance indicators routinely collected to monitor these trends are often a composite of multiple pathogens. For example, ‘influenza-like illness’ — routinely monitored as a proxy for influenza infections — is a symptom definition that could be caused by a wide range of pathogens, including multiple subtypes of influenza, SARS-CoV-2, and RSV. Inferred trends from such composite time series may not reflect the trends of any one of the component pathogens, each of which can exhibit distinct dynamics. Although many surveillance systems routinely test a subset of individuals contributing to a surveillance indicator — providing information on the relative contribution of the component pathogens — trends may be obscured by time-varying testing rates or substantial noise in the observation process. Here we develop a general statistical framework for inferring temporal trends of multiple pathogens from routinely collected surveillance data. We demonstrate its application to three different surveillance systems covering multiple pathogens (influenza, SARS-CoV-2, dengue), locations (Australia, Singapore, USA, Taiwan, UK), scenarios (seasonal epidemics, non-seasonal epidemics, pandemic emergence), and temporal reporting resolutions (weekly, daily). This methodology is applicable to a wide range of pathogens and surveillance systems.

## Introduction

The time-series of incidence of new infections through time is an important metric for monitoring the spread of an infectious disease and the impact of interventions. The true time series of infection incidence cannot be measured — to do so would require that all infections (including asymptomatic and mild infections) were detected and their timing known. Instead, epidemic dynamics are inferred using the time series of quantities more amenable to surveillance that are expected to correlate with infection levels (e.g. the daily number of symptomatic cases) [1]. However, these surveillance indicators often reflect trends in a composite of multiple pathogens, variants, subtype, or serotypes that each exhibit their own distinct dynamics [2–4] (hereafter composite time series). For example, the surveillance of respiratory pathogens such as influenza relies on the monitoring of ‘influenza-like illness’ (ILI) and ‘acute respiratory infections’ (ARI). Both surveillance indicators are based on individuals reporting to healthcare with symptoms, which could be caused by (infection with) a wide range of pathogens (e.g., SARS-CoV-2, influenza, RSV) including multiple influenza types and subtypes [5,6]. Surveillance indicators for SARS-CoV-2, such as laboratory confirmed cases, have often been composed of multiple highly (genetically) distinct variants [7].

Inferring epidemic dynamics from composite time series, under the assumption that they correlate with the infection levels of only a single pathogen, can lead to biased results. For example, during a period of variant replacement (e.g., the replacement of the SARS-CoV-2 Alpha variant during the emergence of the Delta variant) the incidence of one variant may be increasing, while that of the other declines. Analysis of the combined signal would obscure the risk of a future resurgence in infections. Disease prediction could be improved — enhancing the effectiveness of public health responses — if the epidemic dynamics of individual pathogens could be disentangled [5] from a composite time-series.

Many countries collect additional data on component pathogens that contribute to the composite time series. For example: individuals presenting with ILI may be tested for influenza with positive cases typed and subtyped; SARS-CoV-2 cases may be sequenced and the variant determined; and laboratory confirmed dengue virus infections may be serotyped. This data is currently underutilised in public health surveillance reports; the time series of component data is regularly presented separately to the composite time series as a stacked bar chart [8,9]. Although this may allow visual inspection of trends in the pathogen represented by the bottom bar, it obscures trends of the other ‘stacked’ pathogens. Additionally, virological testing rates can vary over time obscuring temporal trends in infections, and when testing rates are low, substantial noise will be present in daily or even weekly data, hampering visual interpretation. If statistical methods existed for combining composite time series with data on the component pathogens, the individual dynamics of each pathogen could be inferred and visualised. For example, during the COVID-19 pandemic models were developed to infer the relative-growth trends of novel variants of concern, particularly during their emergence [3,10,11]. Additionally, some mechanistic models have been fitted to time series of ILI and testing data simultaneously [12,13]. However, these models have often been disease and context specific and so may not be generalisable.

Here we develop a general methodological framework for inferring the epidemic dynamics of multiple pathogens, variants, subtypes or serotypes from routinely collected pathogen surveillance data. We take a non-parametric Bayesian approach, modelling the individual epidemic curves of each pathogen as distinct stochastic processes (random-walks or penalised-splines). We fit the individual epidemic curves to data describing the relative contribution of each pathogen to the component time series. Simultaneously, we fit the sum of all curves to the composite time series. We demonstrate the applicability of our model across multiple settings and pathogens by: inferring the dynamics of influenza subtypes in Australia, Singapore, and the United States of America (USA) from 2012–2024; quantifying the role of SARS-CoV-2 variants in driving epidemic dynamics from 2020–2023 in the United Kingdom (UK); and describing the contribution of dengue virus serotypes to dengue dynamics in Taiwan (province of China) from 2006–2016 and during a recent large outbreak in late-2023.

## Methods

### Statistical modelling framework

We extend an existing statistical modelling framework for inferring the trends of up to two pathogens from a composite time series [3,14], as described in full in the Supplementary Methods. The model takes as input:

1. a time series of count data (e.g., the daily number of influenza-like illness cases) where it is expected that the signal is composed of multiple pathogens (i.e. the composite time series); and
2. a time series of count data describing the number of tests positive for each component pathogen of interest and the number of negative tests where applicable (i.e. the component time series).

The model then estimates the expected value of the time series (e.g., a smoothed trend in the daily number of cases accounting for noise) for each individual pathogen. From the modelled trend for each pathogen we can estimate the pathogen’s growth rate and effective reproduction number, *R(t)*, over time [14]. By comparing the modelled trends of two pathogens we can estimate the relative growth rate advantage (additive) and *R(t)* advantage (multiplicative) over time [3]. All code is publicly available (see Data and code availability).

### Data

#### Influenza-like illness

Influenza-like illness data was retrieved from the World Health Organization’s Global Influenza Programme [15]. We collated weekly data for Australia, Singapore, and the USA from the week starting 2 January 2012 to the week starting 25 December 2023 inclusive. The data described: (1) the weekly number of cases of influenza-like illness; and (2) the weekly number of specimens positive for influenza by subtype (and the number of negative tests). We grouped the influenza specimens into: influenza A subtype not determined; influenza A H3N2; influenza A H1N1 (influenza A H1N1, influenza A H1N1pdm09); influenza B (influenza B Yamagata, influenza B Victoria, influenza B lineage not determined); and negative tests (SFig 1–3). We fit the statistical model to these weekly data assuming that the ILI case time series was composed of cases of influenza A H3N2, influenza A H1N1, influenza B, and unknown pathogens. Unknown pathogens described the proportion of ILI that was unattributable to influenza A or influenza B infection (i.e., negative tests).

#### SARS-CoV-2 cases

SARS-CoV-2 case data for the United Kingdom was retrieved from the United Kingdom Health Security Agency’s data dashboard [16]. We downloaded the daily SARS-CoV-2 case numbers by specimen date for the United Kingdom from 2020–2022.

Additionally, we downloaded data describing the daily number of variants detected by collection date [7] (SFig 4). The data classified all sequences based on ‘major lineage calls’. We considered 11 groupings of the variants based on their major lineage calls: B.1.177; B.1.1.7 (Alpha variant); B.1.617.2 (Delta variant); BA.1 (Omicron BA.1 variant); BA.2 (Omicron BA.2 variant), BA.4 (Omicron BA.4 variant); BA.5 (Omicron BA.5 variant); BA.2.75 (Omicron BA.2.75 variant); BQ.1 (Omicron BQ.1 variant); XBB (a recombinant of omicron sub-variants); and other. The other category included all lineages with a designation not consistent with any of the major lineage calls. The lineages grouped in the other category are entirely wild type lineages until late-March 2021, and after March 2021 are almost entirely recombinant variants (excluding XBB). Note that B.1.177 was a variant that became dominant in Europe but was not estimated to have any transmission advantage [17].

We considered the period of time from 23 September 2020 to 31 December 2022. The earliest date considered was chosen as the first in which there were 10 samples in the variant dataset. We fit the statistical model to this daily data assuming that the SARS-CoV-2 case time series was composed of cases from all 11 variant groupings. Note that in the periods before a variant had emerged no detections of that variant would be present in the data, and so modelled estimates of the variant’s activity would be close to zero (i.e. <<1 modelled cases).

#### Dengue cases

Dengue case data was retrieved from the Taiwan Centers for Disease Control [18]. We downloaded the daily number of dengue fever cases detected. For a subset of cases, the serotype of infection (1, 2, 3 or 4) was known. We aggregated the data into: (1) a time series describing the daily number of dengue cases by onset date; and (2) a time series describing the daily number of cases for which the serotype was determined, by serotype (SFig 5).

We considered the period of time from 1 April 2006 to 31 March 2016, and the period of time from 1 April 2023 to 31 March 2024. We fit the statistical model to each period of daily data separately assuming that the Dengue case time series contained contributions from the four dengue serotypes. We considered periods running from April to March so that we only include full dengue seasons (dengue activity was typically at a minimum around April). We considered the 2023 season in isolation (see Supplementary Methods) as there was very limited dengue activity from April 2016 to April 2023.

## Results

### Temporal trends in influenza

We estimated the long-term dynamics of influenza subtypes in Australia, Singapore and the USA (Fig 1) from 2012 to 2023 using weekly ILI case numbers and data describing influenza testing/subtyping (i.e. the weekly number of tests determining influenza infection and subtype for a subset of ILI cases). In the USA and Australia, there were distinct seasonal peaks of ILI during (their respective) winter months. Inferred influenza subtype dynamics also exhibited distinct seasonal peaks with similar peak timings to ILI (Fig 1, SFig 6, SFig 7). Peaks in influenza activity were sharper compared to ILI; influenza epidemic activity lasted between 3 and 6 months, with little to no activity between seasons. In contrast, ILI activity was observed all year round with increases in ILI activity often occurring months before any (inferred) increases in influenza. In some influenza seasons, epidemic activity was only observed for a single influenza subtype (e.g. 2012–2013, 2013–2014, 2014–2015 for the USA), but in many seasons there were multiple overlapping influenza epidemics. During these multi-subtype seasons, there was a high degree of synchrony in modelled estimates of epidemic onset and peak timing between influenza subtypes. In Singapore, the timing of epidemics of each influenza subtype was far more variable (compared to the USA and Australia), often with sustained levels of cases between epidemic waves (e.g., H3N2 2016–2018). There were also periods in Singapore with simultaneous epidemic activity of multiple influenza subtypes (e.g., 2016–2020).

**Figure 1:**
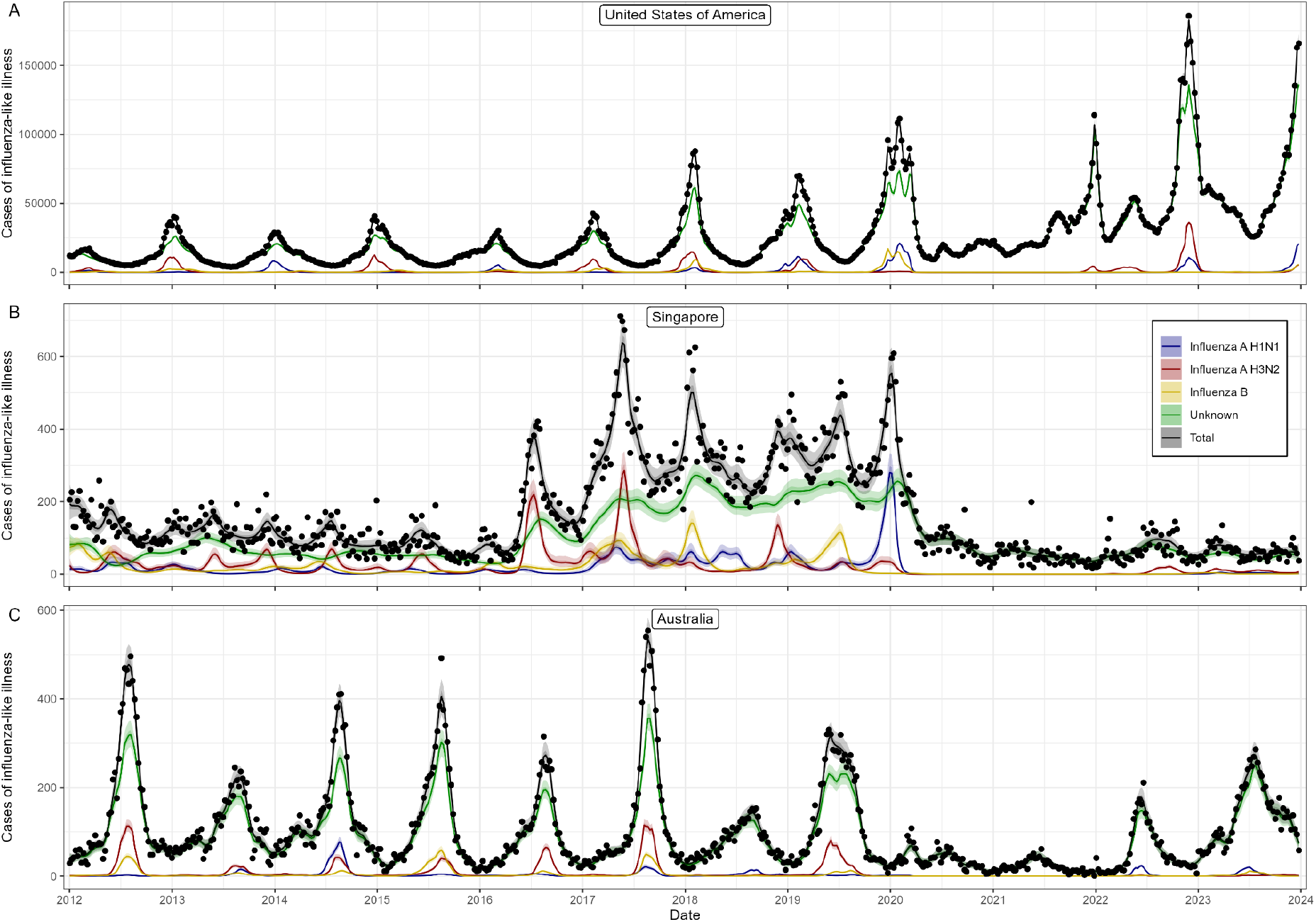
Temporal trends in influenza-like illness and influenza subtypes in the USA, Australia and Singapore. Modelled weekly number of cases attributable to influenza A H3N2 (red), influenza A H1N1 (blue), influenza B (yellow), and not attributable to influenza, ‘unknown’, (green). Weekly number of cases of influenza-like illness (black points) and modelled total number of cases of influenza-like illness (black line). All modelled estimates are shown with median (line) and central 50% (dark shaded region) and 95% (light shaded region) credible intervals.

A high proportion of ILI cases were not attributable to influenza infections (unknown aetiological agent) in all three countries. The proportion of ILI attributable to influenza infections was greatest (relatively consistently) in Singapore (SFig 8), where the influenza positive proportion regularly reached greater than 60% (maximum value of 86%) at the peak of influenza activity. In contrast, in Australia and the USA the proportion of ILI attributable to influenza infections regularly peaked at values between 25% and 35% (maximum value of 40% for Australia and 38% for the USA), even in years when modelled influenza activity was very different (e.g., compare 2015–2016 influenza season in USA to other seasons in SFig 6 and SFig 8). For much of 2020 and 2021 (during the SARS-CoV-2 pandemic), almost all ILI cases were unattributable to influenza infections across all three countries. Following the pandemic period, the proportion of ILI attributable to influenza infections has been lower (on average) in all three countries relative to the pre-pandemic period.

When using lower sampling rates for influenza testing/subtyping than occurred in practice (SFig 9–11), our modelled estimates of subtype dynamics display an increased degree of uncertainty (wider credible intervals) with some features of the epidemic curves — identified at higher sampling rates — obscured. For example, in Singapore from late-2017 to early-2018, at a sampling rate of 20 tests/week a single broad peak of influenza H3N2 was inferred, whereas at higher sampling rates (i.e., 50 tests/week max) a double peak was identified. There was a large degree of agreement between modelled estimates of the total ILI and ILI not attributable to influenza infections (and conversely ILI attributable to influenza), made at all sampling rates considered.

### Temporal trends of SARS-CoV-2 variants

We estimated the dynamics of competing SARS-CoV-2 variants in the United Kingdom (Fig 2) from 23 September 2020 to 31 December 2022 using daily case numbers and the daily number of variants detected (from a subset of cases that underwent sequencing). We captured the dynamics of several periods of variant replacement as novel variants emerged. In November 2020, the Alpha variant (B.1.1.7) emerged with a relatively higher estimated growth rate than existing wildtype variants and the B.1.177 lineage, and Alpha became the sole variant circulating by mid-March 2021. By late-2022, there was a greater variant diversity with multiple variants circulating in the population simultaneously. Omicron BA.1 exhibited the fastest growth rate — on 29 November 2021 (the first day Omicron BA.1 was detected in the UK in the dataset) the growth rate was estimated to be 0.36 (95% CrI: 0.28, 0.45) corresponding to a doubling time of 1.92 (95% CrI: 1.55, 2.47) days. The growth rate of BA.1 then rapidly decreased, becoming less than 0 (the threshold for epidemic decline) only a month later. The replacement of Delta with Omicron BA.1 also reflected the highest growth rate advantage of one variant over the previously dominant variant (SFig 12). Similarly, this period of replacement reflected the highest multiplicative *R(t)* advantage, even when estimated under the assumption that the generation interval declined (SFig 12–13) with each successive variant (see Supplementary Methods).

**Figure 2:**
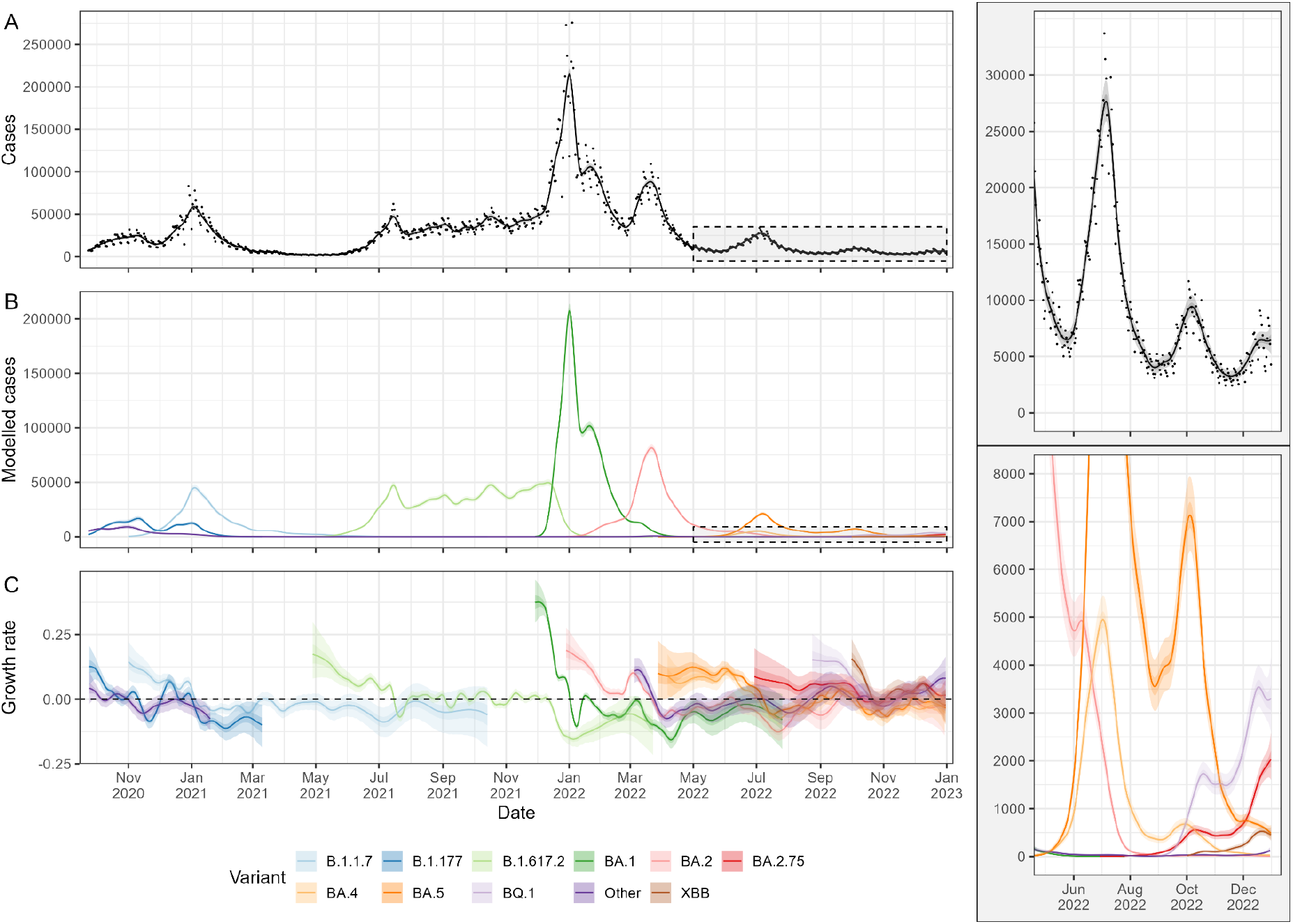
Temporal trends of SARS-CoV-2 variants in the UK. (A). Daily number of cases of SARS-CoV-2 (black points) and modelled total number of cases of SARS-CoV-2 (black line). Shaded box is magnified in the top-right panel. (B) Modelled daily number of cases attributable to each SARS-CoV-2 variant considered (coloured). Shaded box is magnified in the bottom-right panel. (C) Daily epidemic growth rate inferred for each pathogen. The dashed black line highlights an epidemic growth rate of 0 (the threshold for epidemic growth or decline). Estimates for the growth rate of each variant are only shown for times after/before their first/last detection in the UK. Estimates for the growth rate of ‘Other’ are shown up to 19 January 2021 (when the rolling 7 day average first dropped below one) and then shown after 5 March 2022 (when the rolling 7 day average next exceeded one). All lineages categorised as other prior to 19 January 2021 were wild type strains. All estimates are shown with median (line) and central 50% (dark shaded region) and 95% (light shaded region) credible intervals.

Our model can identify increasing components in a composite time series (e.g. SARS-CoV-2 cases), even when the composite time series overall is not increasing. We estimated SARS-CoV-2 case dynamics at six different timepoints (each a week apart) from May 2021–June 2021 (Fig 3) during the emergence of the Delta variant (the first time point was the final day of the week in which the Delta variant was first recorded in the dataset). We compared estimates of growth rates incorporating the component time series (the daily number of each variant detected) and using only the composite time series. When variant data was included in the model, we estimated that Delta cases were likely increasing, although with high uncertainty, at the first two time points, and by the third time point we estimated that Delta cases were increasing with a high degree of certainty (95% CrI for growth rate was greater than 0). In contrast, when only the composite time series was included, there was no clear signal of SARS-CoV-2 cases increasing until the fifth time-point (i.e. five weeks after the Delta variant was first recorded in the dataset) and the overall case growth rate estimated was less than the inferred growth rate of the Delta variant.

**Figure 3:**
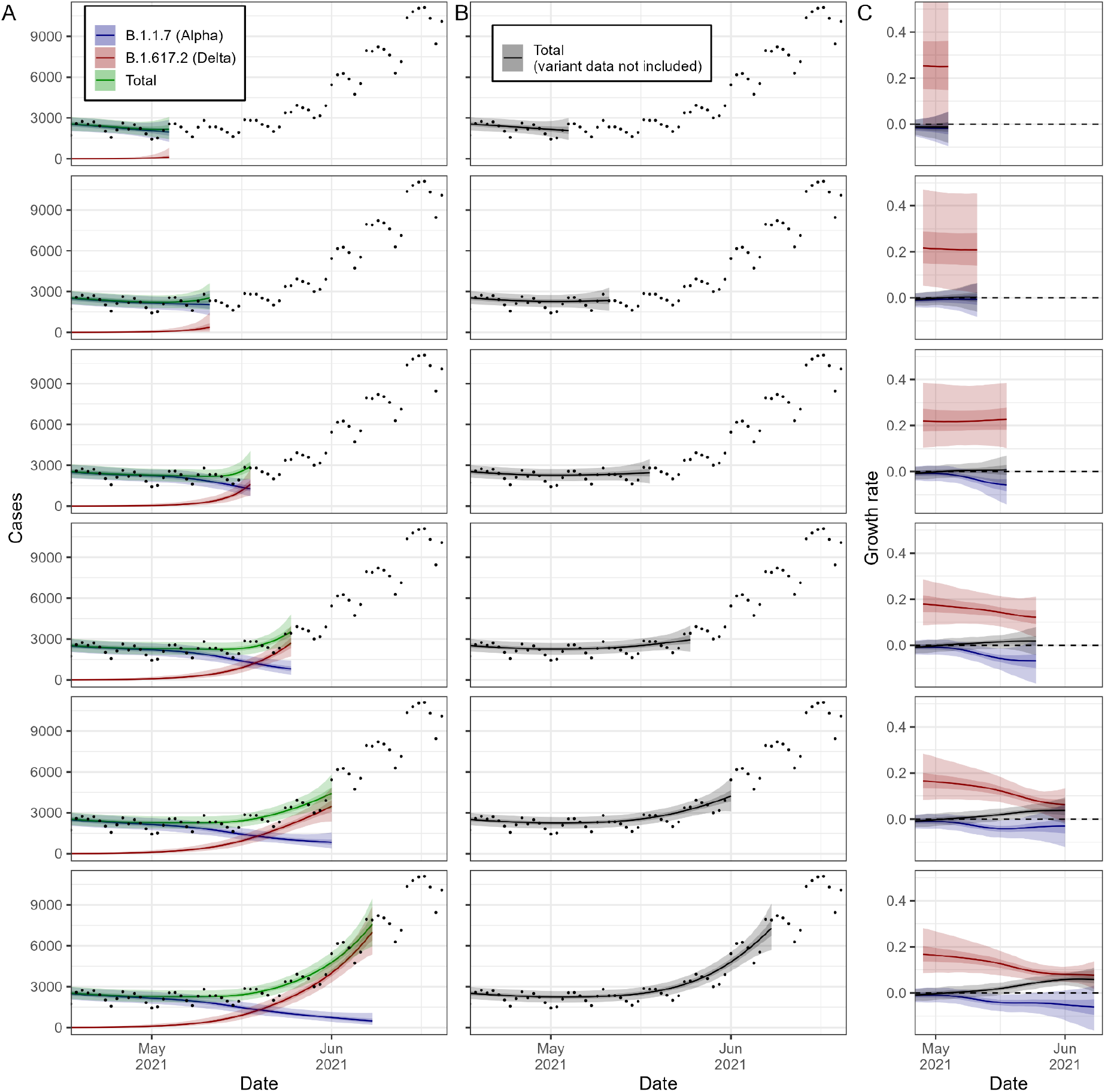
Competing SARS-CoV-2 variant dynamics during the introduction of the Delta variant in the UK. (A) Daily number of cases of SARS-CoV-2 (points) and modelled cases of SARS-CoV-2 attributable to the Alpha variant (Blue), attributable to the Delta variant (Red) and overall (‘Total’, Green). (B) Daily number of cases of SARS-CoV-2 (points) and modelled total cases of SARS-CoV-2 (black) using a model that does not consider the dynamics of different variants (i.e. variant data is not included in the model). (C) Daily epidemic growth rates inferred for the Delta variant (Red), the Alpha variant (Blue), and for all SARS-CoV-2 cases not accounting for competing variant dynamics (from the modelled estimates in B). Estimates for the growth rate of the Delta variant are only shown for times after the first Delta variant had been detected. All estimates are shown with median (line) and central 50% (dark shaded region) and 95% (light shaded region) credible intervals. Models were fit at six different timepoints (see SFig.4) reflecting 1-6 weeks following the first detected Delta variant in the UK).

### Temporal trends of dengue serotypes

We estimated the multi-season dynamics of dengue serotypes in Taiwan (province of China) (Fig 4, Fig 5) using daily dengue (confirmed) case numbers and serotyping data (i.e. the confirmed serotype of infection for a subset of cases) from 2006 to 2016 and 2023 to 2024. There was a seasonal structure to the estimated epidemic dynamics (Fig 4, Fig 5) with limited epidemic activity from January–June each year, and epidemic peaks occurring in approximately October–November and in some years a second peak in July–August (e.g., in 2007). We estimated the proportion of dengue cases caused by each serotype for each season (SFig 14). In most seasons our estimates were consistent with a crude approach (calculating the proportion from the raw serotyping data) that did not account for different dynamics between serotypes, but there were significant differences in 2014 and 2015. The dengue dynamics for a single season were often dominated by a single serotype that was responsible for the majority of cases. The main exception was the 2010 season in which all four dengue serotypes contributed substantially, and to a lesser extent the 2012 season. Even for seasons where we estimated a dominant serotype, there was still limited circulation of at least one, and at times two, other serotype(s) (e.g., in 2011).

**Figure 4:**
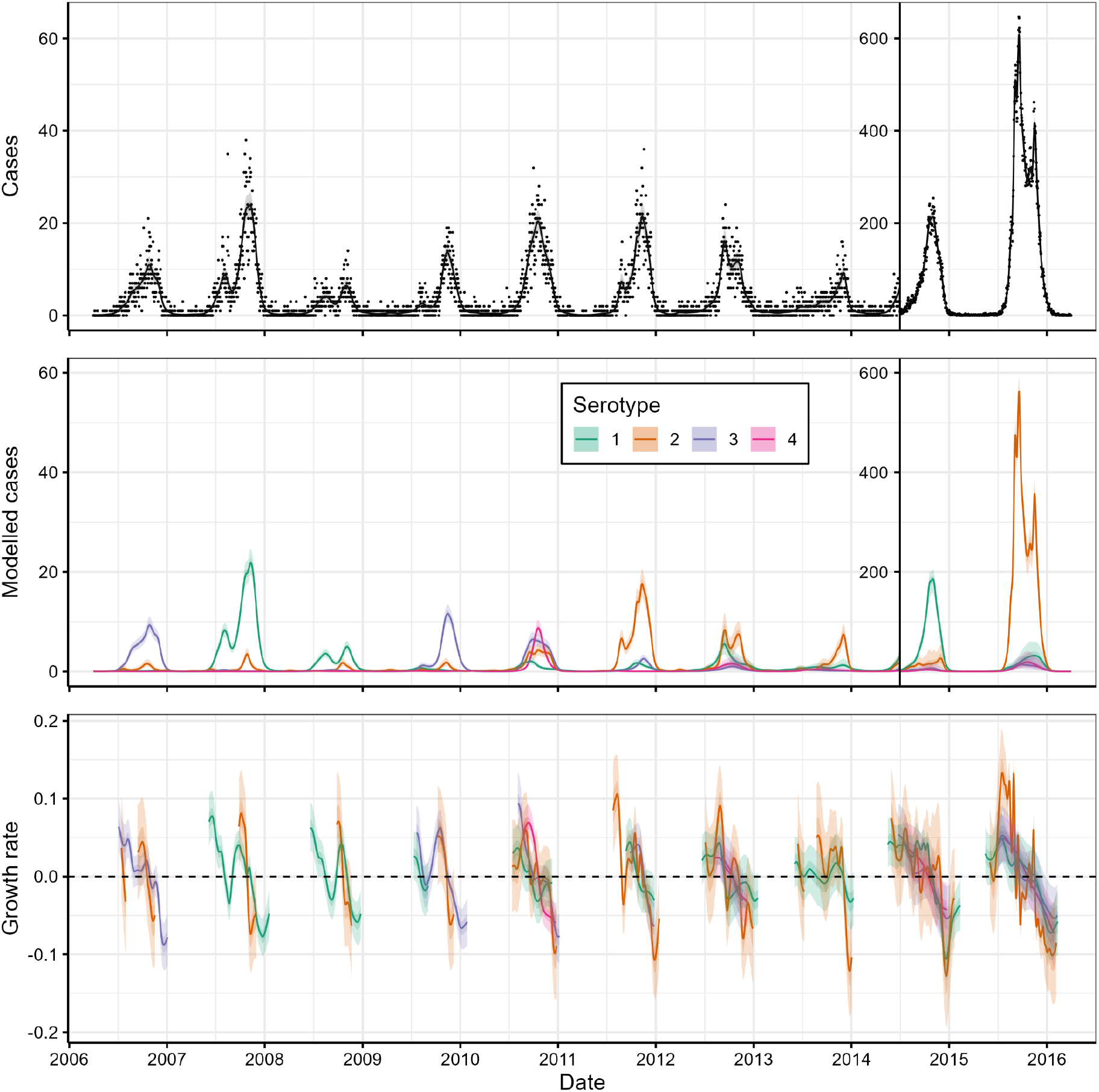
Temporal trends of dengue serotypes in Taiwan (province of China) for the 2006-2015 dengue seasons. (Top panel). Daily number of cases of dengue (black points) and modelled total number of cases of dengue (black line). The y-axis is scaled by a factor of 10 for 1 April 2014 onwards. (Middle panel) Modelled daily number of cases attributable to each dengue serotype (coloured). The y-axis is scaled by a factor of 10 for 1 April 2014 onwards. (Bottom panel) Daily epidemic growth rate inferred for each serotype. Estimates of the growth rate for a serotype are only shown when the modelled serotype case numbers were greater than 0.5. All estimates are shown with median (line) and central 50% (dark shaded region) and 95% (light shaded region) credible intervals.

**Figure 5:**
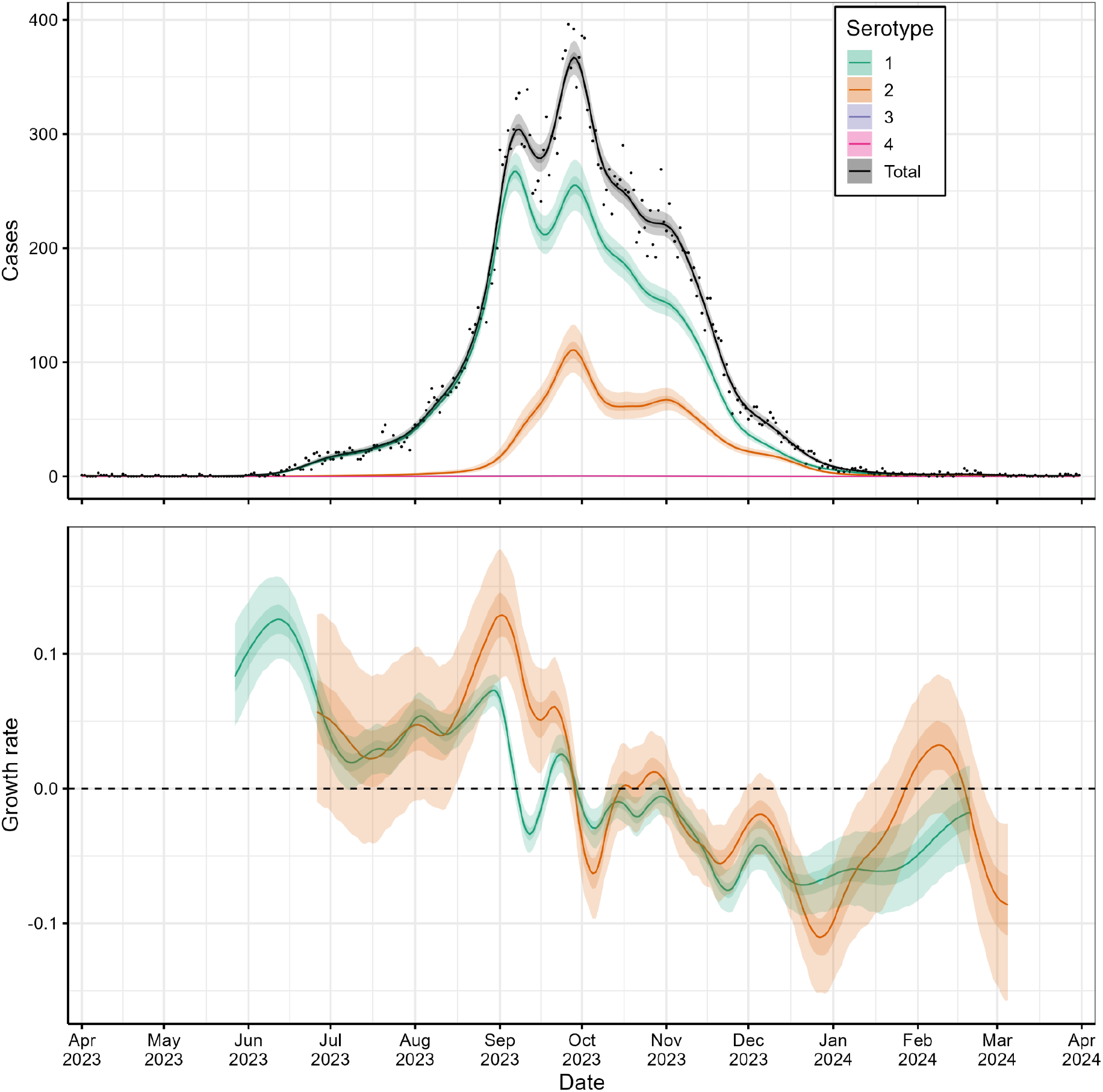
Temporal trends of dengue serotypes in Taiwan (province of China) for the 2023 dengue season. (Top panel). Daily number of cases of dengue (black points) and modelled total number of cases of dengue (black line) and cases attributable to each dengue serotype (coloured). (Bottom panel) Daily epidemic growth rate inferred for each serotype. Estimates of the growth rate for a serotype are only shown when the modelled serotype case numbers were greater than 0.5. All estimates are shown with median (line) and central 50% (dark shaded region) and 95% (light shaded region) credible intervals.

## Discussion

We have developed a method for inferring the epidemic dynamics of multiple pathogens, variants, subtypes or serotypes from routinely collected surveillance data. We have demonstrated its application to three different surveillance systems, each of which produce data on a composite time series (e.g., daily number of cases) and component time series (e.g., daily number of laboratory confirmed infections by pathogen). The systems considered cover multiple pathogens (influenza, SARS-CoV-2, dengue), locations (Australia, Singapore, USA, Taiwan, UK), scenarios (seasonal epidemics, non-seasonal epidemics, pandemic emergence) and data structures (weekly, daily, daily with day-of-the-week effects).

Routine surveillance of influenza predominantly relies on the symptom-based surveillance of influenza-like illness. We have demonstrated that a significant proportion of ILI is not attributable to influenza infections and have estimated the true underlying dynamics of each influenza subtype. The quantity estimated by our model is equivalent to the ‘subtype-specific ILI+’ [19], a surveillance indicator(s) that is crudely calculated by multiplying the number of ILI cases by the proportion of tests positive for a specific subtype. The subtype-specific ILI+ has previously been suggested to be more representative of the overall and subtype-specific influenza dynamics compared to ILI [1]. However, trends in crude estimates of subtype-specific ILI+ may be obscured (limiting its utility) when there is noise in the time series of ILI (the composite time series) and/or the time series describing influenza testing and subtyping data (the component time series). Our statistical model can estimate the expected smooth trends (and credible intervals) in the subtype-specific ILI+ accounting for noise in the observation processes. Improved estimation of the underlying dynamics of influenza infections (overall and by subtype) can help refine estimates of influenza season onset, which is important for optimising the timing of vaccination campaigns. In all three settings investigated, we observed that increases in ILI cases can occur notably earlier than actual increases in influenza infections, suggesting that ILI is a poor indicator for determining influenza season onset.

Influenza-like illness is caused by a wide range of non-influenza pathogens with similar symptom profiles. As one would expect, the proportion of ILI attributable to influenza appeared to have decreased following the emergence of SARS-CoV-2; the continued circulation of (ILI causing) SARS-CoV-2 will likely continue to influence trends in the time series of ILI into the future [6]. The time series of ILI has likely always been biased as an influenza surveillance indicator due to the circulation of other respiratory pathogens such as RSV, rhinovirus, and parainfluenza [20], but now those biases may have changed due to the emergence of SARS-CoV-2. If a subset of ILI cases were tested for a wide array of pathogens through virological testing (as opposed to just testing for influenza and subtyping) the positive proportion of many more ILI-causing pathogens (including SARS-CoV-2) could be measured [4,13] and underlying trends inferred using the methods we have presented.

Even when all cases in a time series are confirmed infections (as opposed to those with a symptomatic diagnosis) the overall signal detected may be a composition of multiple signals. For example, we demonstrated that the composite time series of detected dengue and SARS-CoV-2 infections concealed distinct serotype and variant dynamics respectively. Similarly, if a composite time series of laboratory confirmed influenza infections was used as a surveillance indicator for influenza, it would obscure distinct epidemic dynamics of influenza subtypes. Disentangling these distinct dynamics can be critical for predicting future epidemic trends. For example, patterns of population immunity may be dependent on the population’s past exposure levels to each pathogen [21,22]. Additionally, disentangling each pathogen’s dynamics can be important for assessing the effectiveness of past interventions —pharmaceutical (e.g., antivirals, vaccines) and non-pharmaceutical (e.g., school closures) —against each individual pathogen so that their effectiveness at later times (when a different mixture of pathogens may be circulating) can be anticipated.

Improved knowledge of the individual dynamics of pathogens contributing to a composite time series can improve estimation of current trends [5] (i.e. real-time analysis). In general, if current trends continue into the future, then the composite time series will be dominated by the pathogen with the largest growth rate at the current time point. Outputs from our model can be used to estimate the growth rates of all component pathogens and therefore predict which pathogens are increasing at the fastest rate, and so which pathogens are likely to dominate the composite time series in the future. We have retrospectively demonstrated how the model might have been used for real-time analysis during the emergence of the SARS-CoV-2 Delta variant in the United Kingdom. While trends in modelled Delta cases suggested an increasing component in the SARS-CoV-2 case time series, increasing cases would not have been predicted (or identified) until five weeks after the first Delta sequence was isolated if sequencing data were not included in the analysis. Our estimates of current growth rates (i.e. estimates up to the most recent day) appeared to be robust to the addition of future data points with credible intervals overlapping with estimates made using additional data points. However, a formal performance evaluation is needed to further establish the model’s utility as a tool for real-time analysis.

Our model has limitations. The model assumes that infection incidence increases/decreases exponentially over the short-term; this is an appropriate assumption for many pathogens that exhibit exponential growth in the early phase of an epidemic but may not be appropriate for pathogens, such as measles, that have been observed to grow sub-exponentially [23]. Our model infers pathogen-specific epidemic dynamics at the population-level over an entire region. However, there may be differences in epidemic dynamics between sub-regions and sub-populations (e.g., by age group). The model in its current form could be fit to data for a single sub-region or sub-population in isolation; in future the model could be extended to infer trends in pathogen-specific epidemic dynamics stratified across sub-groups simultaneously allowing for common parameters (e.g., overdispersion in case data) to be estimated using all available data, reducing uncertainty in modelled estimates [24]. Finally, the model can only infer trends that exist in the data. If the data are not a reasonable correlate of the underlying epidemic dynamics, then our modelled estimates will not reflect the true epidemic dynamics. The symptomatic- and case-based surveillance data systems used in this paper have previously been shown to be biased by (for example) circulating pathogens with similar symptom profiles, and changes in healthcare-seeking or test-seeking behaviour of the general population through time [1,25]. There is limited data available without such biases and so we are unable to validate our estimates against the true values. In the future, a simulation-estimation study could be used for further testing and validation of the methodology.

Virological testing is a highly important component of pathogen surveillance. However, to date there has been limited routine analyses that integrate virological testing data with analysis of other composite epidemic time-series. One exception is analyses performed during the SARS-CoV-2 pandemic that incorporated genomic sequencing data into time series analysis of cases and infection prevalence (which motivated the approach presented here [3]). We have developed a statistical method to better utilise routine surveillance data and demonstrated its utility for surveillance systems measuring influenza-like illness, SARS-CoV-2 and dengue cases, but it likely has wider applications. For example, RSV strains are separated into two groups (A and B) and measuring the dynamics of each individually may be important for assessing if pharmaceuticals (such as maternal vaccines or monoclonal antibodies) are equally effective against infection with both groups [26]. Similarly, Mpox is composed of two distinct clades (I and II) each with two distinct subclades (Ia, Ib, IIa, and IIb). Measuring the dynamics of each (sub)clade individually will be crucial for estimating the spread and transmission advantage, if present, of the newly emergent clade Ib [27]. In general, our methodology can enhance the epidemic intelligence obtained from these routine surveillance data, improving our understanding of the dynamics of multiple pathogens, and ultimately improving public health responses.

## Data Availability

All the underlying time series data is publicly available. The publicly available time series data is also provided at the same location as the code https://github.com/acefa-hubs/EpiStrainDynamics/tree/preprint (Zenodo DOI: https://doi.org/10.5281/zenodo.14015867).

## Data and code availability

The code that produced this analysis is publicly available at: https://github.com/acefa-hubs/EpiStrainDynamics/tree/preprint (Zenodo DOI: https://doi.org/10.5281/zenodo.14015867). All the underlying data is publicly available, but is also provided at the same location as the code.

