## Supplementary Materials for "Inferring temporal trends of multiple pathogens, variants, subtypes or serotypes from routine surveillance data"

**Supplementary methods**

***Statistical modelling framework***

*Overview*

We extend an existing statistical modelling framework for inferring the trends of up to two pathogens [1,2]. The mixed-effects Bayesian P-spline model (described in [1]) can infer the temporal trends of two competing variants from a time series of infection prevalence data (daily number of positive and negative tests) and data describing the variant identified for a subset of sequenced positive specimens. We have extended this model to: handle any number of pathogens; fit to time series data of counts (e.g. daily number of cases); incorporate influenza testing data in which the subtype for influenza A samples may be undetermined; account for day-of-the-week effects in daily data; include options for fitting penalised splines or random walks; support additional (optional) correlation structures in the parameters describing the smoothness of the penalised splines (or random walks); and include additional (optional) sources of noise in the observation process.

*A smoothed model for expected cases*

A general model for the expected number of cases can be written in terms of a smoothed function:

$$g(\pi(t)) \; = \; s(t).$$

Here, $\pi(t)$ is the expected case time series at time $t$, $s(t)$ is the value of a smoothed function at time $t$, and $g$ is the link function, which we take as the logarithmic function for count data (e.g. a case time series). If we expect the composite time series to be composed of signals from multiple pathogens we can extend this general model to include a smoothed term for each component pathogen ($i$):

$$g(\pi_i(t)) \; = \; s_i(t).$$

Where, $s_i(t)$, is now the value of a smoothed function describing component pathogen $i$ at time $t$, and $\pi_i(t)$ is the modelled case time series for each component pathogen $i$ at time $t$ (i.e. the contribution of component pathogen $i$ to the overall composite time series) . We consider two different smoothed functions: the penalised-spline and the random-walk (see below).

*Penalised-spline model*

A penalised-spline is a linear combination of $N$ basis-splines of degree $n$:

$$s_i(t) \; = \; \sum_{j=1}^{N} b_{i,j} B_{j,n}(t).$$

Here $b_{i,j}$ are the set of $N$ basis-spline coefficients for pathogen $i$, and $B_{j,n}(t)$ is the value of the $j^{th}$ basis-spline of order $n$ as a function of time $t$.

Basis-splines are defined recursively over a system of non-decreasing (temporally) equidistant knots: $t_1$, ...., $t_{N+n-2}$. The first-order ($n = 1$) basis splines are defined as:

$$B_{j,1}(t) = 1 \quad , t_j < t \le t_{j+1}$$

$$= 0 \quad , otherwise.$$

Higher-order basis splines are then defined as:

$$B_{j,n}(t) = w_{j,n} B_{j,n-1} + (1 - w_{j+1,n}) B_{j+1,n-1} ,$$

where

$$w_{j,n} = \frac{t-t_j}{t_{j+n-1}-t_j} \quad , t_j \neq t_{j+n-1}$$

$$= 0 \quad , otherwise.$$

When defining the system of knots we specify the number of days between adjacent knots. The number of days between adjacent knots was assumed to be five days for all analyses performed in this paper. This has been demonstrated to be an appropriate choice when fitting to SARS-CoV-2 infection prevalence time series [2] as it captures most of the dynamical effects (fine enough temporal resolution) while not being too computationally expensive — the main benefit of using penalised-splines over the random-walk model. We also define the system of knots to extend beyond the beginning and end of the period the data covers. This is to prevent edge effects in the model; $n$ (the order of basis splines) knots on each edge will not have the same form as interior knots. We consider basis splines of order $n$=3 for all analyses and so the system of knots is always defined to extend by three additional boundary knots before and after the period the data covers.

*Random-walk model*

A random-walk assumes that for each time-step there is a new parameter:

$$s_i(t) = b_i(t).$$

Where $b_i(t)$ takes a new value at each time-step according to a probability distribution (see second-order random-walk prior distribution below). This is analogous to a penalised-spline of order one with daily knots. Henceforth we will refer to the coefficients of the penalised-spline ($b_{i,j}$) and the random-walk ($b_i(t)$) interchangeably as $b_{i,j}$.

*Second-order random-walk prior distribution*

In order to avoid overfitting of the model we must assume a relationship between coefficients of the smooth functions $s_i(t)$ (penalised spline or random-walk). We assume a second order random-walk prior distribution on the coefficients, which has previously been used when modelling epidemic dynamics [2]. The second-order random-walk prior can be written as:

$$b_{i,n} = 2b_{i,n-1} - b_{i,n-2} + u_{i,n},$$

where

$u_{i,n} \sim Normal(0,\ \tau_i)$.

This prior distribution penalises changes in the first derivative of the smooth function. The amount that changes in the first derivative are penalised is set by the parameter(s) $\tau_i$. We consider two different assumptions for the dimensions of $\tau_i$. When fitting the model to data on influenza-like illness and dengue we assume that each pathogen $i$ has its own corresponding $\tau_i$. When fitting the model to data on SARS-CoV-2 we assume a single parameter $\tau$ describes the smoothness of the trends for all SARS-CoV-2 variants. This was assumed due to the many extra parameters that would be required (11 variants considered), and the significant periods of time in which each pathogen was at approximately 0 cases (limited data to infer trends for each pathogen individually). One could also consider a two-dimensional version of $\tau_i$ ($\tau_{ik}$)in which it describes variance and covariance between changes in the first derivative of each pathogen $i$ and each other pathogen $k$. This was not considered in this paper, but is an option for model fitting implemented in the code (see code availability statement).

*Likelihood*

The model is fit to a composite time series of count data (e.g. the number of daily or weekly cases), $C(t)$, and a component time series (of the same temporal resolution) of count data describing the number of samples which were positive for each component pathogen (e.g. the number of influenza A H3N2, influenza A H1N1, influenza B positive tests and the number of negative tests), $P_i(t)$.

We assume that the composite time series is negative-binomially distributed:

$$C(t) \sim Negative\ Binomial(\sum_i \pi_i(t),\ \eta).$$

Where we have defined the negative binomial distribution in terms of its mean, $\mu$, and overdispersion, $\eta$: $NB(\mu, \eta)$. In this formulation of the negative binomial distribution the mean is simply $\mu$, and the variance is $\mu\,(1\ +\ 1/\eta)$. We take the mean (for a given point in time) to be the value of the sum of the modelled time series for each pathogen, and the overdispersion, $\eta$, to be an additional parameter of the model.

We assume that the component time series, describing the number of samples positive for each pathogen, is multinomially distributed:

$$P_i(t) \sim Multinomial(\sum_i P_i(t), \frac{\pi_i(t)}{\sum_i \pi_i(t)}).$$

Where we have defined the the multinomial distribution in terms of the number of trials, $n$, and the probability of each independent outcome, $p_i$: $M(n,\ p_i)$. We take the number of trials to be the total number of samples in our component time series (for a given time point), and the probability of each outcome to be the relative value of each pathogen's modelled time series.

Additionally, influenza-like illness subtyping data includes samples positive for influenza A (unsubtyped), influenza A H3N2, and influenza A H1N1. To infer the dynamics of influenza A H3N2 and influenza A H1N1 separately (as opposed to grouping all influenza A together), we adjust the multinomial likelihood above slightly. Firstly, for the multinomial likelihood we do not consider the modelled time series of influenza A H3N2 and influenza A H1N1 separately, we consider their combined total: $\pi_{infA}(t) = \pi_{H3N2}(t) + \pi_{H1N1}(t)$. We then fit to the data ($P_i(t)$) grouping all influenza A samples together (influenza A subtype not determined, influenza A H3N2 and influenza A H1N1). Secondly, we assume that the data describing the number of each influenza A subtype (H3N2 and H1N1), for the subset in which influenza A subtype is determined, is binomially distributed:

$$P_{H3N2}(t) \sim Binomial(P_{H3n2}(t) + P_{H1N1}(t), \frac{\pi_{H3N2}(t)}{\pi_{H3N2}(t)+\pi_{H1N1}(t)}).$$

Where, $P_{H3N2}(t)$ is the number positive for H3N2, $P_{H1N1}(t)$ is the number positive for H1N1, $\pi_{H3N2}(t)$ is the modelled time series for H3N2, and $\pi_{H1N1}(t)$ is the modelled time series for H1N1.

*Model fitting*

The model was implemented in STAN [3]. Posterior parameter distributions were sampled using the default No-U-Turns Sampler (NUTS) [4]. We ran four chains for 5,000 iterations with burn-in of 1,000 iterations. The likelihood of the model was as defined above. We assumed non-informative constant priors for all parameters (including parameters defined below). We describe the specific model options that were used when fitting to each set of data in supplementary table 1.

*Including day-of-the-week effects*

We can include day-of-the-week effects in the observation process where we expect that testing and/or reporting (and therefore the number of cases in the dataset) changes based on the day of the week. We simply include a simplex (vector that adds to one) parameter of length $k$ ($k = 7$ for a different effect for each day), $\theta_k$, describing the multiplicative day-of-the-week effect in the likelihood function for the composite time series (e.g. case time series):

$$C(t) \sim Negative\ Binomial(k \times \theta_k \times \sum_i \pi_{ik}(t), \eta).$$

Here, $\pi_{ik}(t)$ is the modelled time series for each pathogen $i$, and $k$ simply describes the day of the week classification. We assume a day-of-the-week effect when fitting our model to daily case data for SARS-CoV-2. We assume that there is a different effect for each day of the week (i.e., $k = 7$). It is also possible (and easy to implement in our available code) to group days of the week together; for example one could consider a different effect for weekdays vs weekends ($k = 2$, two states depending on whether the day is a weekday or weekend).

This formulation assumes that the underlying transmission process (i.e. $\pi_i(t)$) does not include a day-of-the-week effect (and so is well described by a smooth function), but that the observation process (i.e. testing) includes the effect.

*Including additional noise between pathogens*

Noise in the data is captured by the overdispersion parameter of the negative binomial distribution (i.e. the parameter $\eta$ from the 'likelihood' section). This assumes that the noise structure is the same for all pathogens. This is likely when most of the noise is due to the observation process (testing and reporting) and not due to stochastic fluctuations (which may not be correlated) of the pathogen's infection dynamics. We also considered including an additional source of noise that allows additional noise that can vary between pathogens.

We assume an additional sampling step. The modelled time series including noise for each pathogen, $\overline{\pi_i(t)}$, is assumed to be gamma distributed:

$$\overline{\pi_i(t)} \sim Gamma(\pi_i(t),\ \phi).$$

Where we have defined the gamma distribution in terms of its mean, $\mu$, and overdispersion, $\phi$. In this formulation of the gamma distribution the mean is simply $\mu$, and the variance is $\mu/\phi$. We take the mean (for a given point in time) to be the modelled time series for each pathogen, and the overdispersion, $\phi$, to be an additional parameter of the model. All likelihoods above are then written instead in terms of the modelled time series including noise ($\overline{\pi_i(t)}$ instead of $\pi_i(t)$).

We fit the model including the additional source of noise to influenza-like illness data for the USA and Australia. There was a high degree of agreement between modelled estimates for each pathogen's time series (SFig 15-16) for the model fit with and without the additional source of noise. Estimated parameters of $\phi$ and $\eta$ suggested that the majority of the noise was in the observation process.

We estimated the ratio of the overall overdispersion due to each source of noise: $\frac{1+1/\eta}{1/\phi}$.

The ratio was 56 (44, 73) for the USA and 169 (148, 195) for Australia.

When fitting to our other case study datasets, the inclusion of the additional noise term introduced convergence problems. This was likely because the origin of the noise in these datasets was often indistinguishable and so the two overdispersion parameters ($\phi$ and $\eta$) were degenerate.

*Inclusion of more informative prior distributions*

The full model (with non-informative constant prior distributions) would not converge when fitting to a single season of dengue data. This was likely due to limited data available over a single season to capture the overdispersion of the negative-binomial distribution, $\eta$, and the smoothness parameters for each serotype's penalised-spline, $\tau_i$.

However, the posterior parameter distributions for $\eta$ and $\tau_i$ (from the model fit to multiple seasons of data) could be incorporated as the prior distributions for $\eta$ and $\tau_i$ when fitting the model to a single season. This can be included in the sampling process as:

$$\eta \sim Normal(\bar{\eta}, Var(\eta))$$

$$\tau_i \sim Normal(\bar{\tau}_i, Var(\tau_i)).$$

Where we have defined the posterior parameter distributions of $\eta$ and $\tau_i$ in terms of their mean ( $\bar{\eta}$ and $\bar{\tau}_i$) and variance ($Var(\eta)$ and $Var(\tau_i)$) and assumed that they are normally distributed.

We estimated the dengue dynamics of the 2014 season and 2015 season separately using the parameter posterior distributions (means and variances) obtained fitting the full model to data from the 2006–2013 seasons as prior distributions. The estimated dengue dynamics for both seasons were consistent with those made fitting the full model to all data from the 2006–2015 seasons (SFig 17). Using this method, we also estimated the dengue dynamics for the 2023 season (that could only be inferred in isolation due to the near zero dengue activity from 2016-2023) using parameter posterior distributions from the model fit to the data from the 2006–2015 seasons.

***Estimating other epidemiological quantities***

Epidemiological quantities, such as the growth rate and effective reproduction number, can be estimated from the smoothed epidemic time-series for each pathogen. For each smoothed time series the epidemiological quantity of interest (see below) can be calculated for each sample in the posterior distribution giving us the posterior distribution of the epidemiological quantity of interest. We report the central and 95% credible intervals of the posterior distribution.

*Growth rate*

The instantaneous growth rate, $r_i(t)$, of a pathogen's epidemic time-series can be estimated at time $t$ using the simple equation:

$$r_i(t) = log(\pi_i(t)) - log(\pi_i(t-1)).$$

As we are using a log link function for modelling the case time series in this paper, this is equivalent to:

$$r_i(t) = s_i(t) - s_i(t-1).$$

The doubling/halving time of an epidemic time-series is simply defined as $log(2)/r_i(t)$

*Growth rate advantage*

The growth rate advantage, $\Delta r_{ij}(t)$, of one pathogen, $i$, over another pathogen, $j$, is simply the additive difference between their growth rates:

$$\Delta r_{ij}(t) = r_i(t) - r_j(t).$$

*Effective reproduction number*

The time-varying reproduction number describes the expected number of secondary infections caused by an individual infected on day $t$ if conditions were to remain the same.

The reproduction number for a pathogen's epidemic time series can be calculated using the equation:

$$R_i(t) = \frac{\pi_i(t)}{\int_0^{\inf} \pi_i(t-\chi) g_i(\chi) d\chi}.$$

Here $g_i(\chi)$ is the generation time distribution of pathogen $i$, and describes the distribution of time between primary infection (individual is infected) and secondary infection (individual infects someone else). Estimates of $R_i(t)$ are sensitive to the assumed generation time distribution [5]. This equation assumes that the modelled epidemic time-series is a reasonable proxy to the infection incidence, in reality the epidemic time-series will likely be lagged and smoother compared to actual infection incidence; this could lead to slightly lagged estimates of $R_i(t)$ that may also be biased when the true value of $R_i(t)$ changes rapidly [6]. Additionally, the epidemic time-series will not account for any changes in the case ascertainment over time, potentially introducing further bias to estimates of $R_i(t)$.

*Multiplicative reproduction number advantage*

The multiplicative reproduction number advantage, $\text{♀} R_{ij}(t)$, of one pathogen, $i$, over another pathogen, $j$, is simply the multiplicative difference of their reproduction numbers:

$$\text{♀} R_{ij}(t) = \frac{R_i(t)}{R_j(t)}.$$

We estimate the multiplicative reproduction number advantages of multiple pairs of SARS-CoV-2 variants: B.1.1.7 (alpha variant) over wild type (lineages classified as "other" prior to 19 January 2021 — 19 January 2021 is the last date in which the rolling 7 day average number of 'other' lineages detected was greater than 7, until 5 March 2022 at which time "other" lineages did not describe wildtype); B.1.617.2 (Delta variant) over B.1.1.7; BA.1 (Omicron BA.1 variant) over B.1.617.2; BA.2 (Omicron BA.2 variant) over BA.1; and BA.5 (Omicron BA.5 variant) over BA.2. We initially estimate all reproduction numbers (and multiplicative advantages) assuming a constant gamma distributed generation time distribution between variants. We then estimated all reproduction numbers assuming successively shortened generation time distributions (reduced means) with similarly shaped distributions (same shape parameter between gamma distributions). See supplementary table 2 for all generation time distributions used (visualised in SFig.13) [7,8].

### Supplementary figures

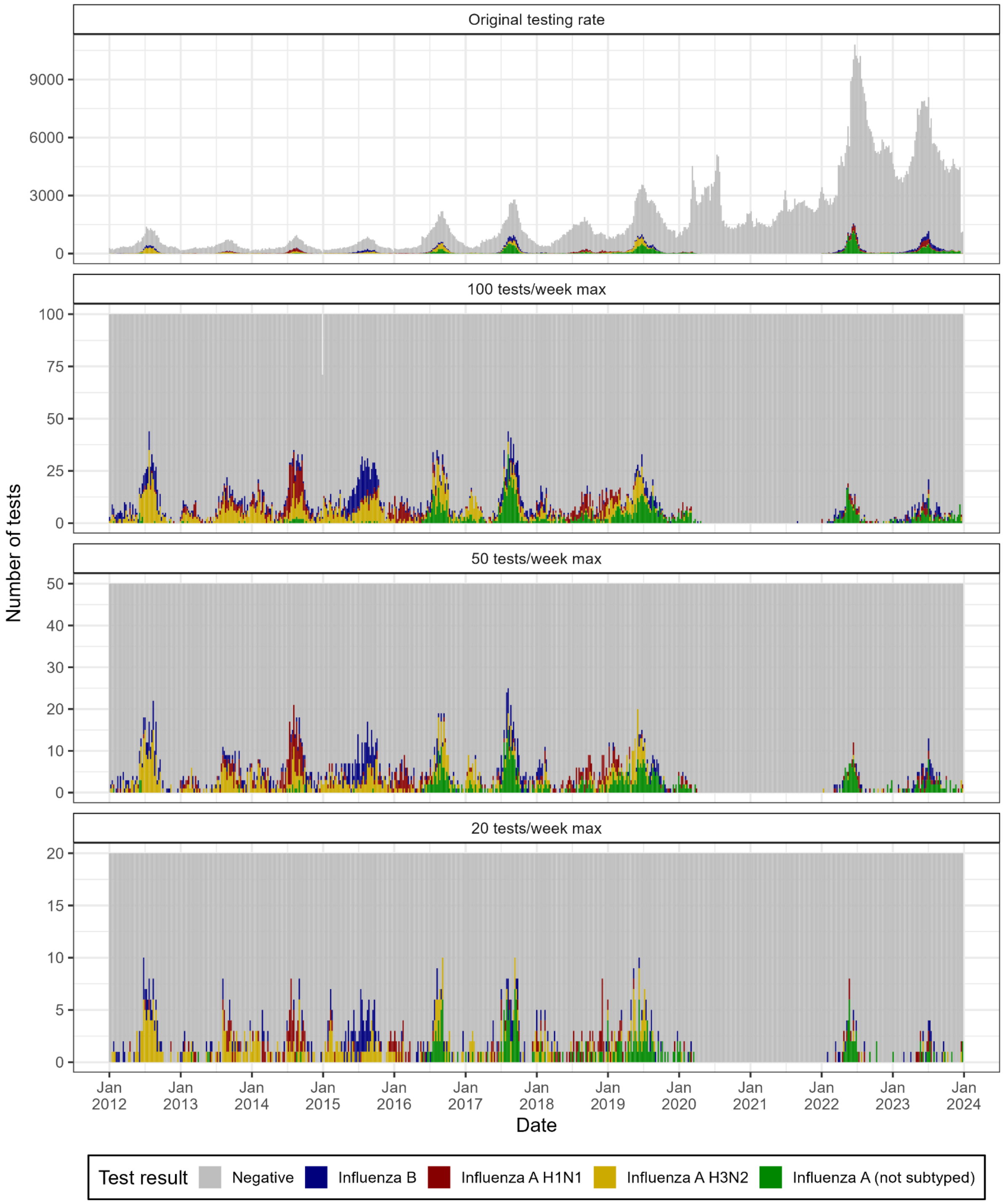


**SFig 1: Influenza-like illness testing and subtyping data for Australia.** Weekly number of influenza-like illness cases that undergo testing and the result of the test. Test results are divided into negative tests (Grey), tests positive for influenza B (Blue), and tests positive for influenza A with subtype: not determined (Green); determined to be H3N2 (Yellow); and determined to be H1N1 (Red). The top panel shows the original testing data used for the main analyses. The bottom three panels show randomly selected subsamples of the original weekly testing data with maximum number of tests per week of 100 (second panel), 50 (third panel) and 20 (fourth panel).

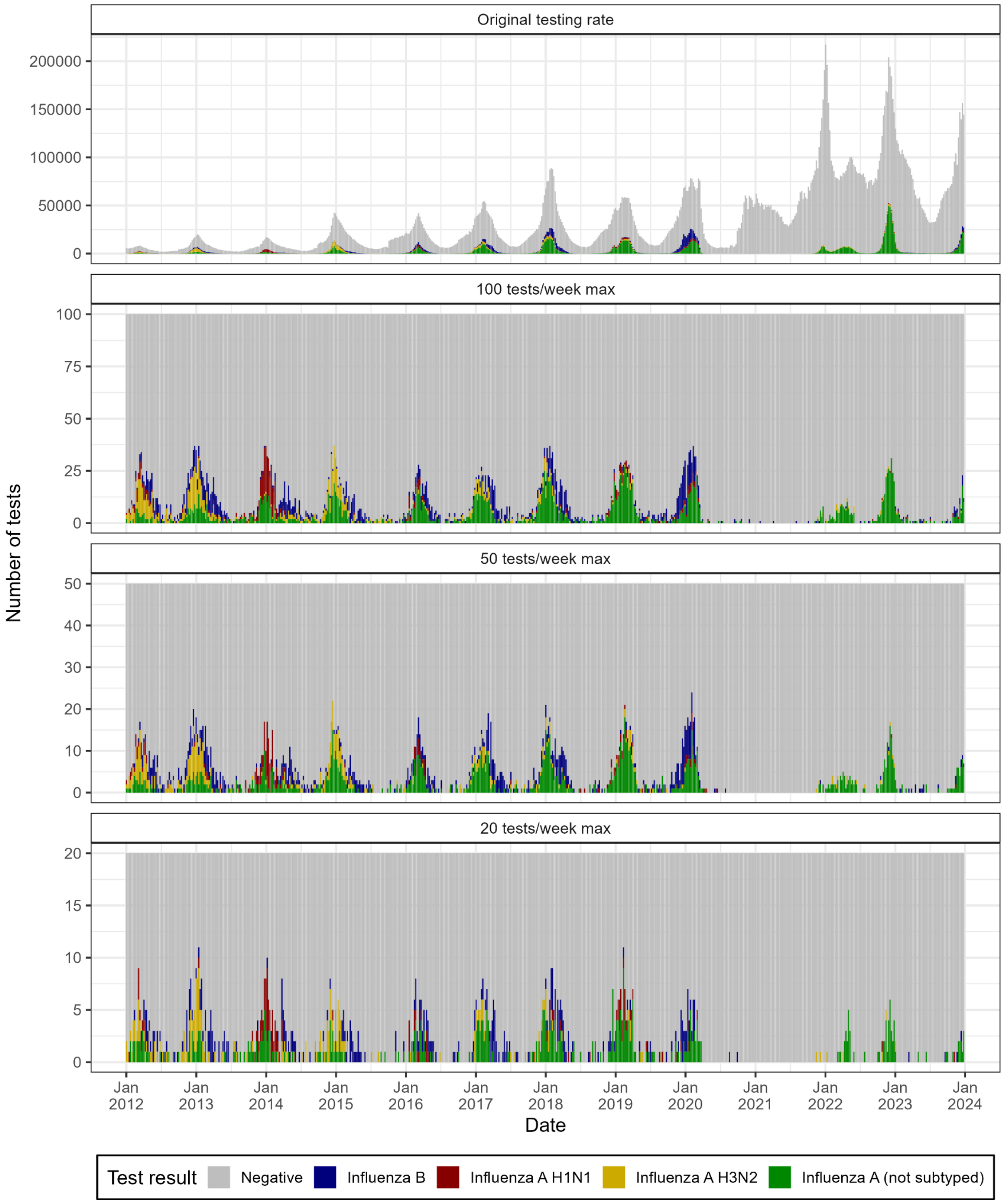


**SFig 2: Influenza-like illness testing and subtyping data for the USA.** Weekly number of influenza-like illness cases that undergo testing and the result of the test. Test results are divided into negative tests (Grey), tests positive for influenza B (Blue), and tests positive for influenza A with subtype: not determined (Green); determined to be H3N2 (Yellow); and determined to be H1N1 (Red). The top panel shows the original testing data used for the main analyses. The bottom three panels show randomly selected subsamples of the original weekly testing data with maximum number of tests per week of 100 (second panel), 50 (third panel) and 20 (fourth panel).

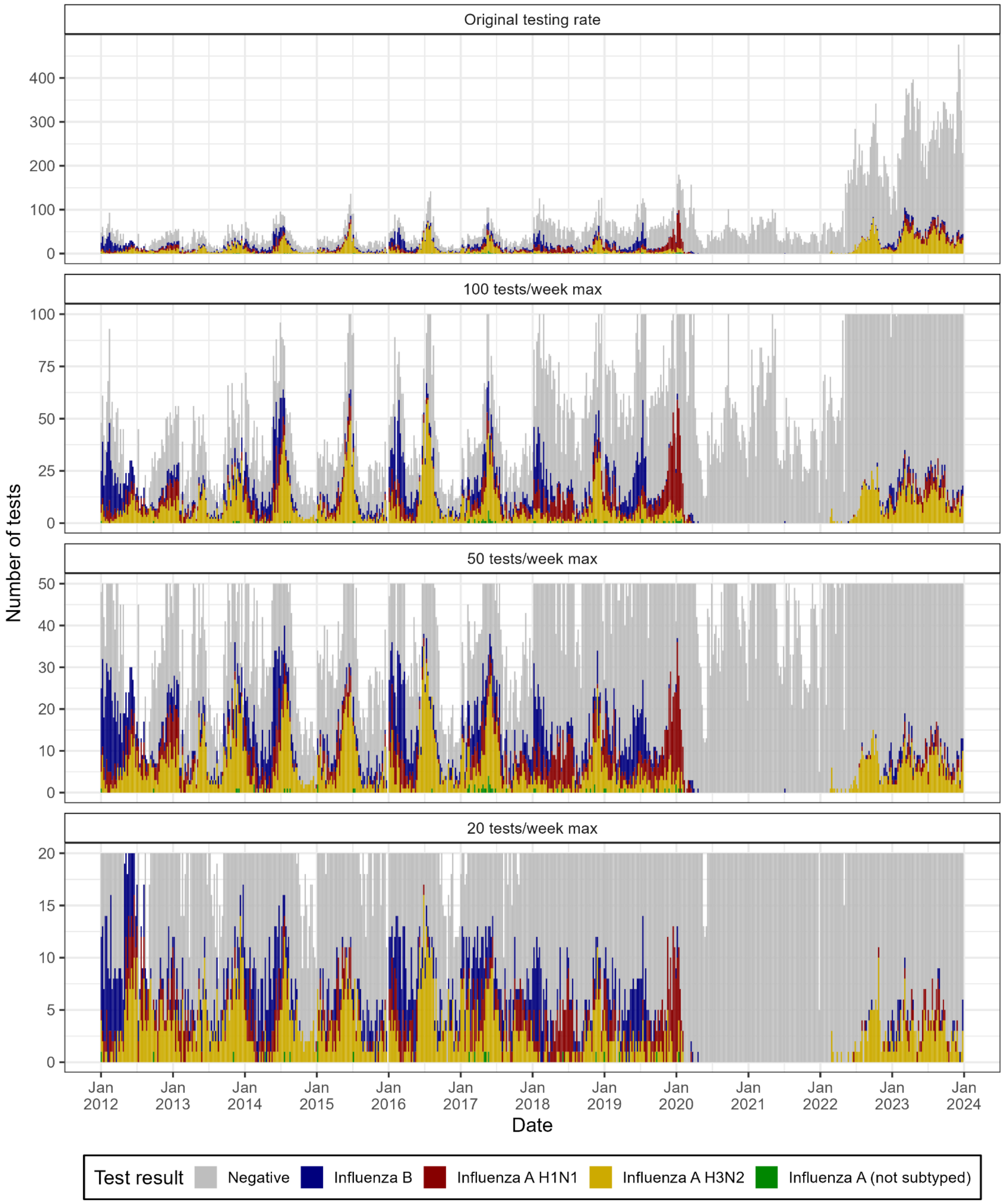


**SFig 3: Influenza-like illness testing and subtyping data for Singapore.** Weekly number of influenza-like illness cases that undergo testing and the result of the test. Test results are divided into negative tests (Grey), tests positive for influenza B (Blue), and tests positive for influenza A with subtype: not determined (Green); determined to be H3N2 (Yellow); and determined to be H1N1 (Red). The top panel shows the original testing data used for the main analyses. The bottom three panels show randomly selected subsamples of the original weekly testing data with maximum number of tests per week of 100 (second panel), 50 (third panel) and 20 (fourth panel).

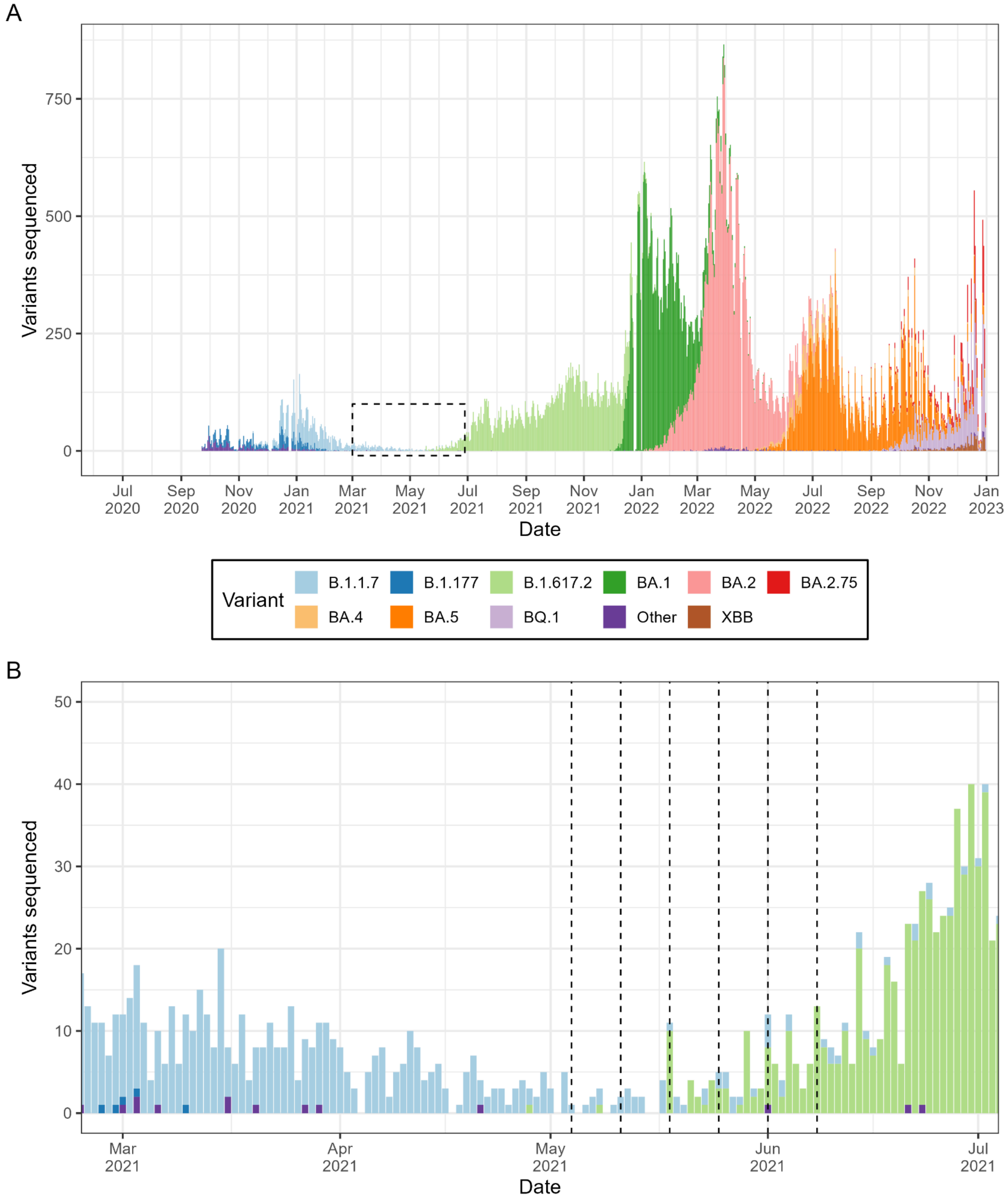


**SFig 4: SARS-CoV-2 variant data for the UK.** (A) Daily number of SARS-CoV-2 variants sequenced in the UK [9]. Sequences were divided into 11 variants (colours) based on groupings of SARS-CoV-2 lineages. (B) A magnified version of the dashed box in panel A during the emergence of the Delta variant. Models presented in Fig.3 were fit to all data up to and including each vertical dashed line. The first dashed line is on 4 May 2021, the final day of the week in which the Delta variant was first detected. The dashed lines are each one week apart.

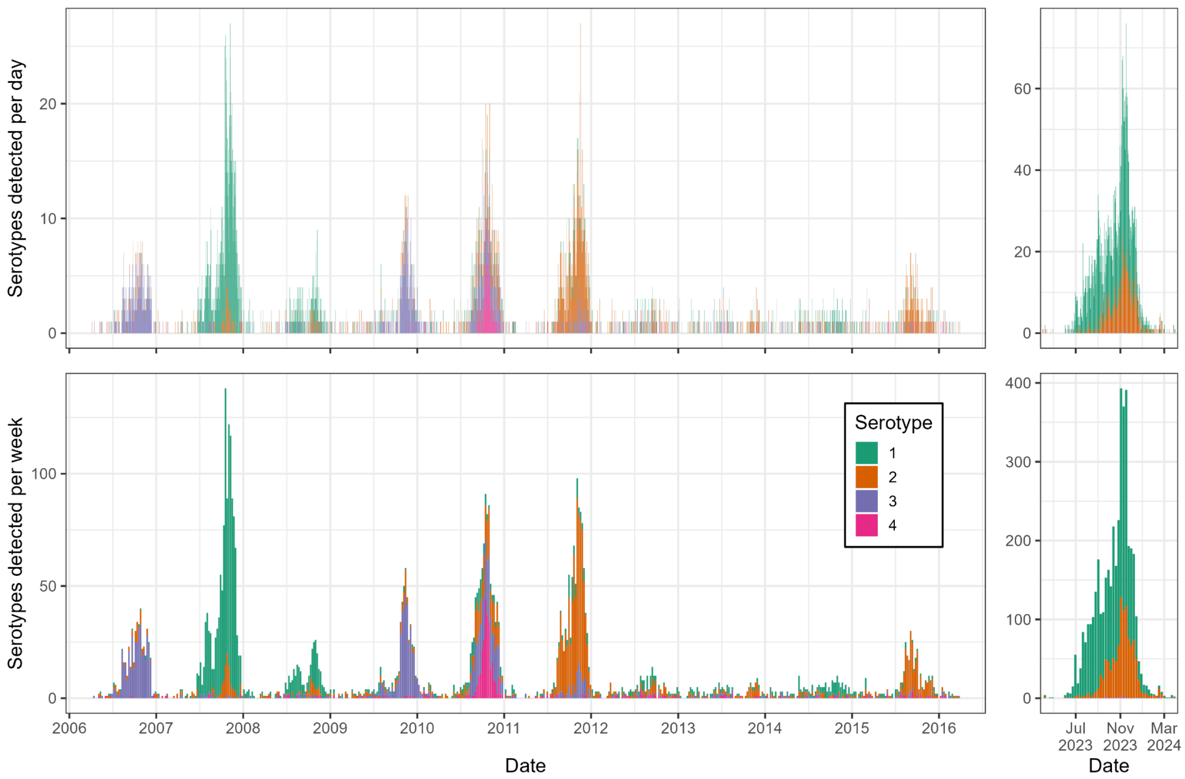


**SFig 5: Dengue serotype data for Taiwan (province of China).** (Top panels) Daily number of dengue serotypes determined in Taiwan (province of China) for the 2006-2015 seasons and the 2023 season. (Bottom panels) Weekly number of dengue serotypes determined in Taiwan (province of China) for the 2006-2015 seasons and the 2023 season. In all analyses daily dengue data was used; we present the weekly numbers only for easier visual interpretation of the raw data.

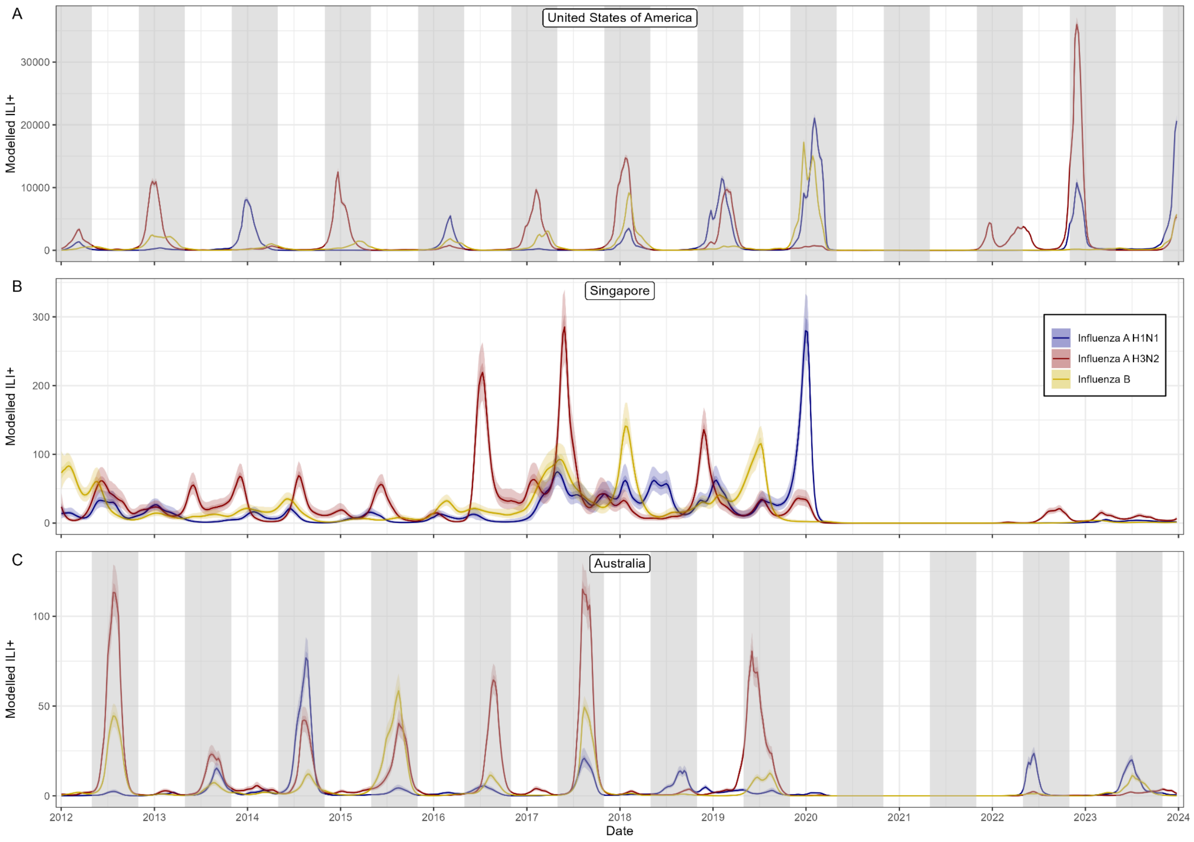


**SFig 6: Temporal trends in influenza subtypes in the USA, Australia and Singapore.** Modelled weekly number of cases attributable to influenza A H3N2 (red), influenza A H1N1 (blue), and influenza B (yellow). All modelled estimates are shown with median (line) and central 50% (dark shaded region) and 95% (light shaded region) credible intervals. Shaded regions represent the 'winter season' in the northern (for the USA) and southern (for Australia) hemisphere. The northern winter season highlighted is from 1 November to 30 April. The southern winter season highlighted is from 1 May to 30 October.

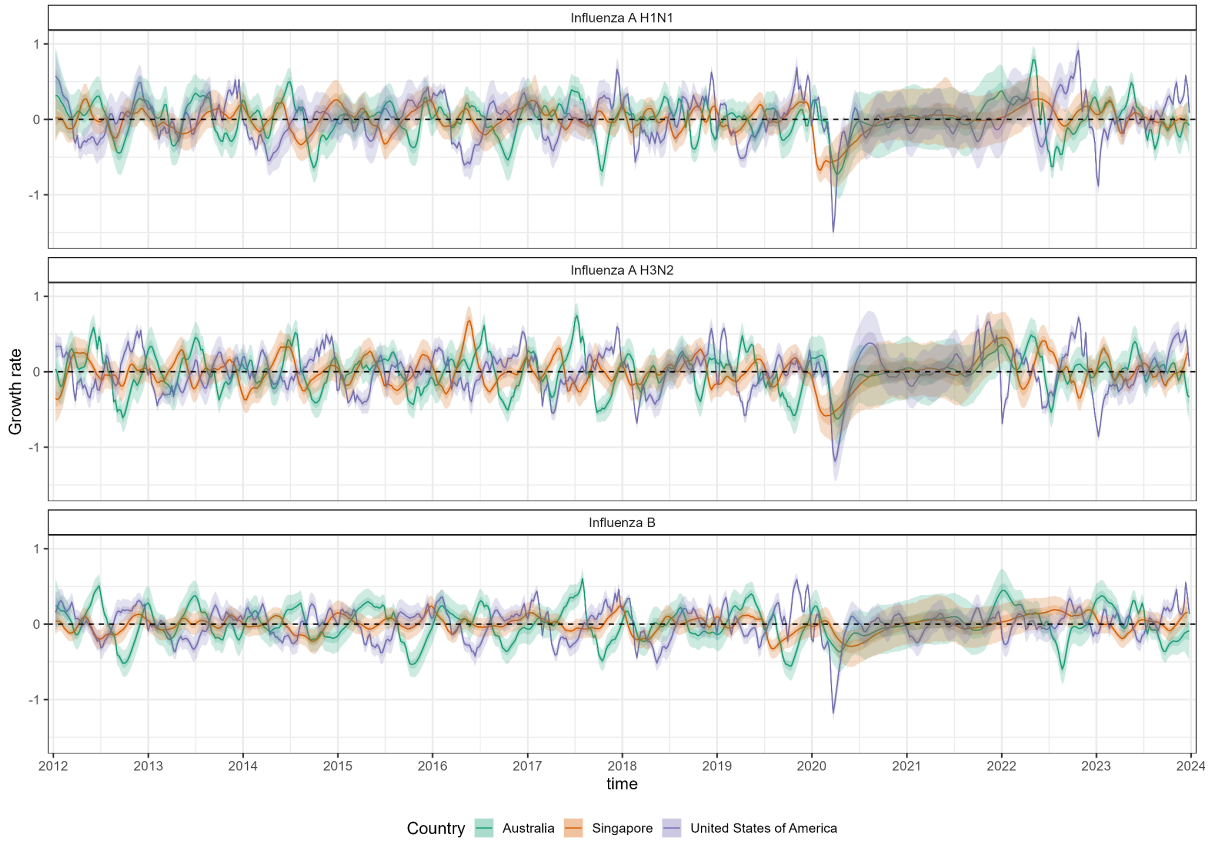


**SFig 7:Temporal trends in the epidemic growth rate of influenza subtypes in the USA, Australia and Singapore.** Weekly epidemic growth rate inferred for each influenza subtype (panels) in Australia (Green), Singapore (Orange), and the USA (Purple). The dashed black line highlights an epidemic growth rate of 0 (the threshold for epidemic growth or decline). All modelled estimates are shown with median (line) and central 50% (dark shaded region) and 95% (light shaded region) credible intervals.

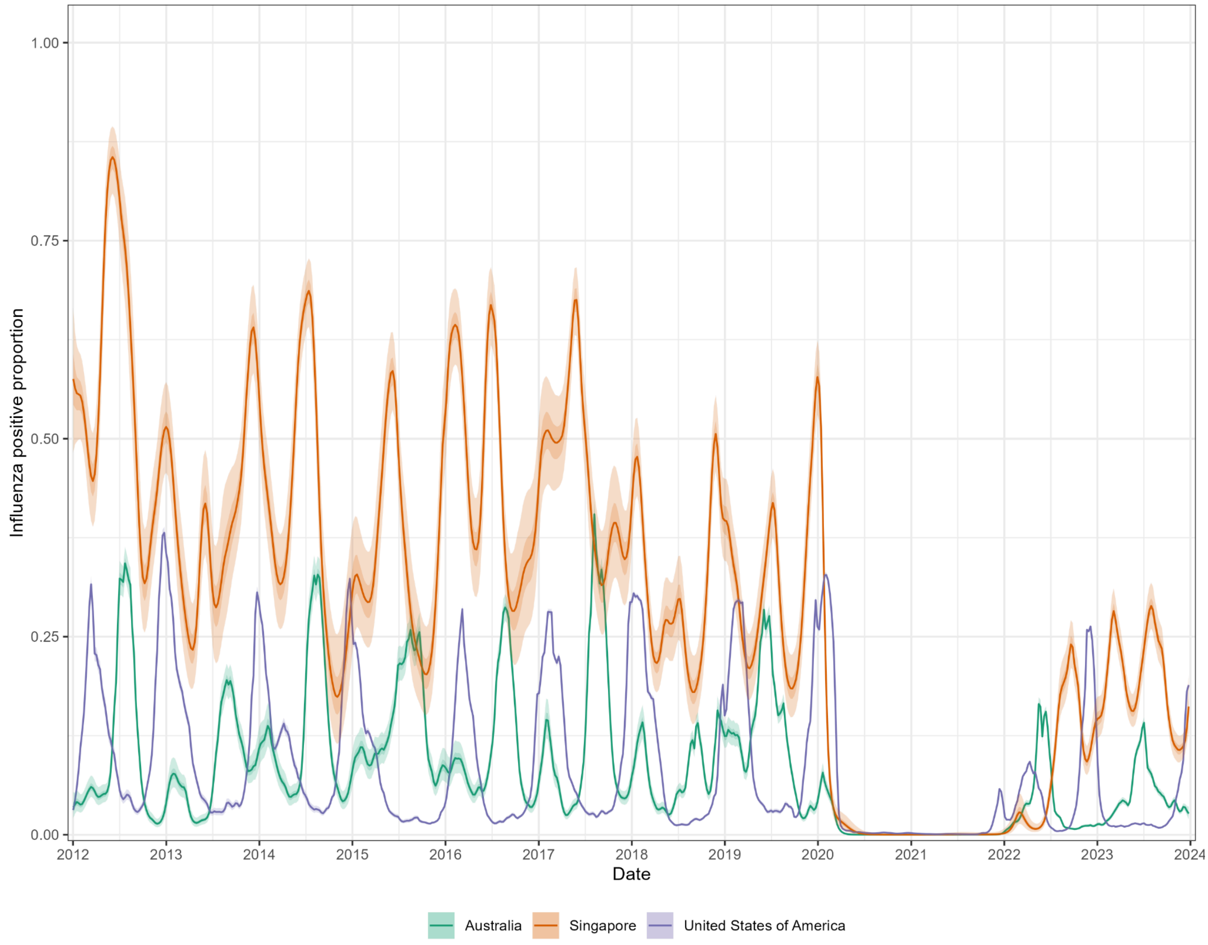


**SFig 8: The influenza positive proportion over time in the USA, Australia and Singapore.** The weekly proportion of cases of ILI that are attributable to an influenza infection for the USA (Purple), Singapore (Orange), and Australia (Green). All modelled estimates are shown with median (line) and central 50% (dark shaded region) and 95% (light shaded region) credible intervals.

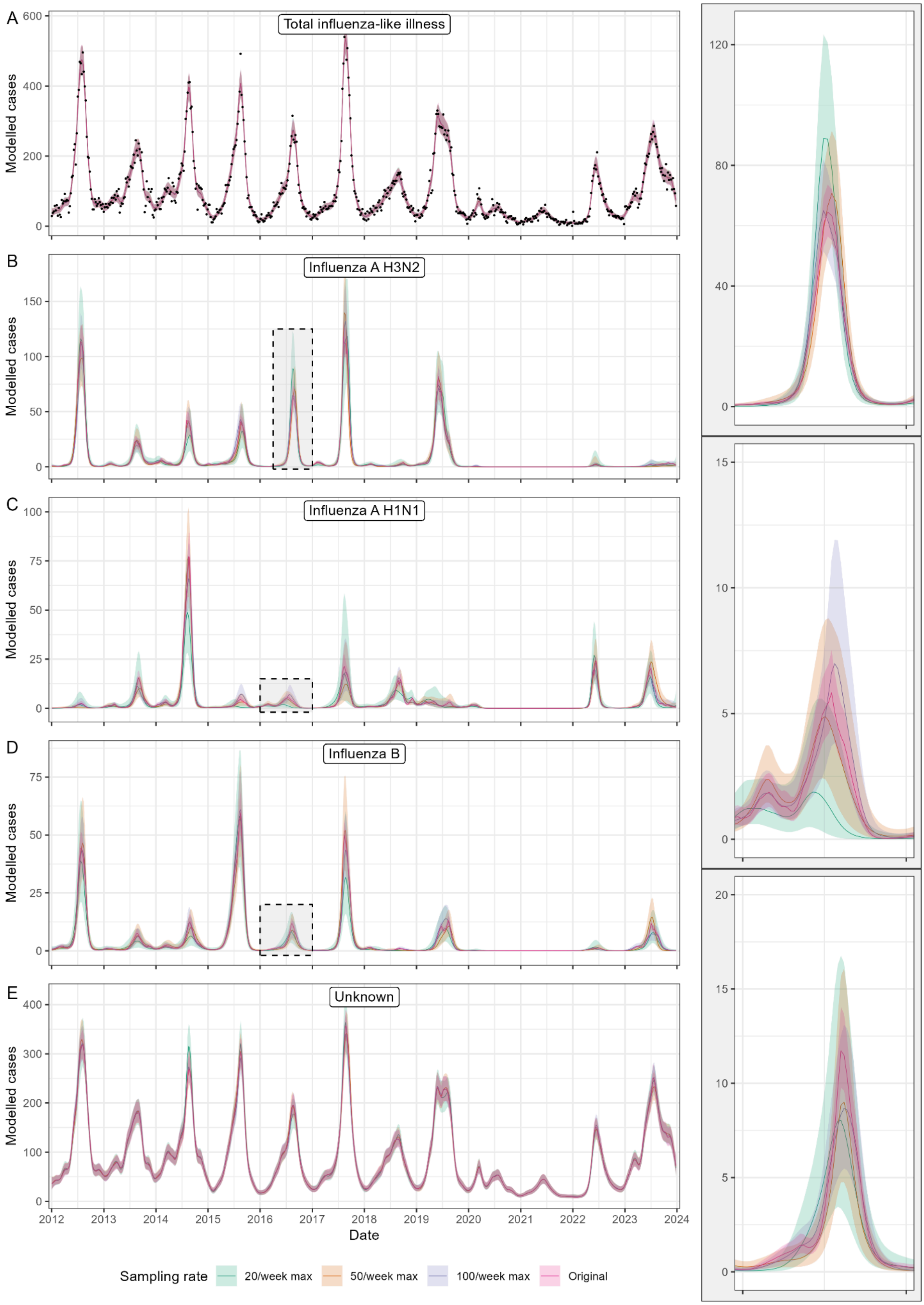


**SFig 9: Trends in influenza-like illness and influenza subtypes in Australia inferred at different testing rates.** (A) Modelled total number of weekly cases of influenza-like illness. (B-E) Modelled weekly number of cases attributable to influenza A H3N2 (B), influenza A

H1N1 (C), influenza B (D), and not attributable to influenza, ‘unknown’, (E). The three shaded boxes are shown magnified in the same order in the right-hand panels. For all panels modelled cases are shown for estimates made assuming cases of ILI were tested at the original sampling rate (Pink) or a maximum of 100 tests (Purple), 50 tests (Orange), or 20 tests (Green) a week. See SFig.1 for a visualisation of the testing data used at each sampling rate. All modelled estimates are shown with median (line) and central 50% (dark shaded region) and 95% (light shaded region) credible intervals.

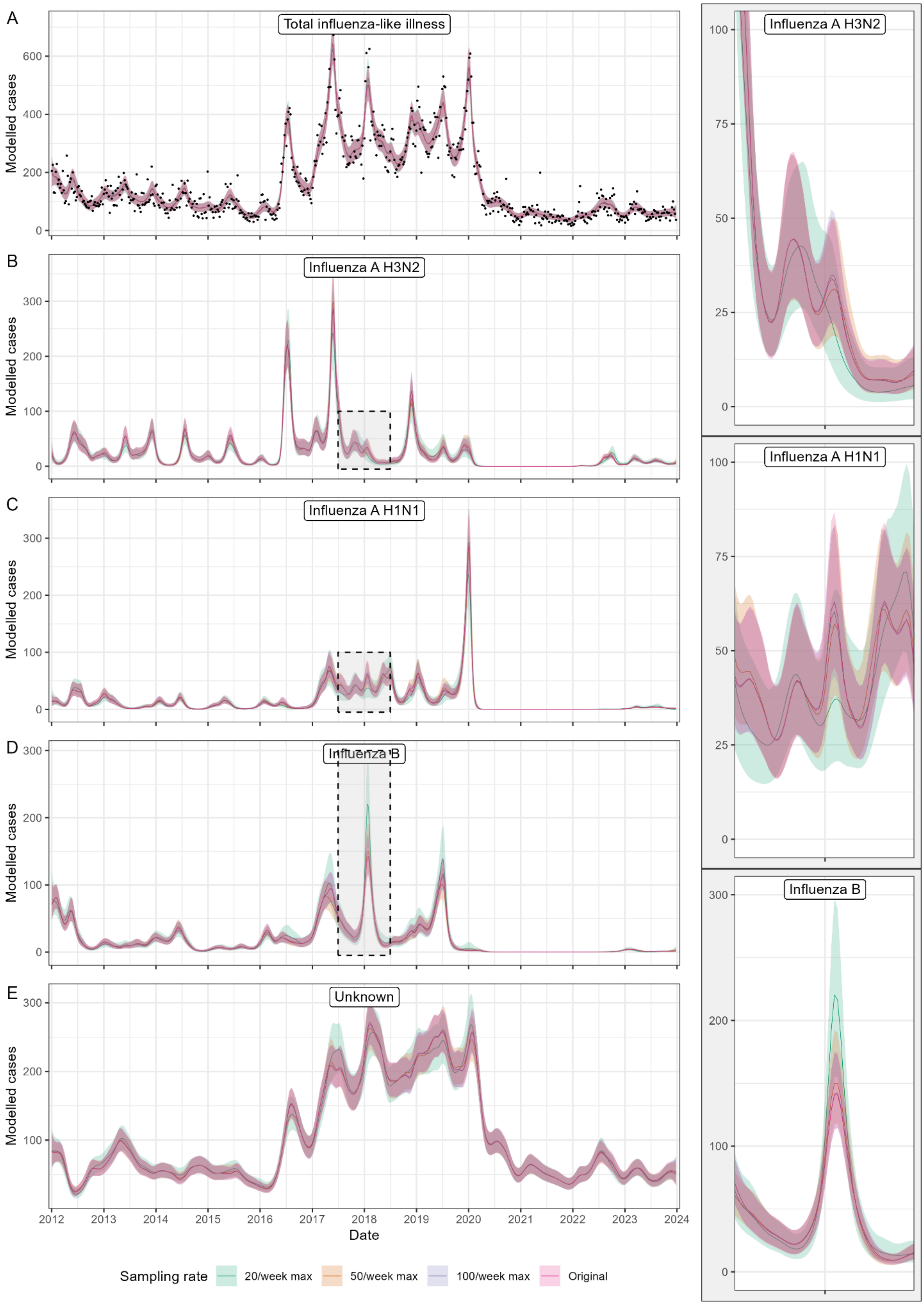


**SFig 10:Trends in influenza-like illness and influenza subtypes in Singapore inferred at different testing rates.** (A) Modelled total number of weekly cases of influenza-like illness. (B-E) Modelled weekly number of cases attributable to influenza A H3N2 (B),

influenza A H1N1 (C), influenza B (D), and not attributable to influenza, ‘unknown’, (E). The three shaded boxes are shown magnified in the same order in the right-hand panels. For all panels modelled cases are shown for estimates made assuming cases of ILI were tested at the original sampling rate (Pink) or a maximum of 100 tests (Purple), 50 tests (Orange), or 20 tests (Green) a week. See SFig.3 for a visualisation of the testing data used at each sampling rate. All modelled estimates are shown with median (line) and central 50% (dark shaded region) and 95% (light shaded region) credible intervals.

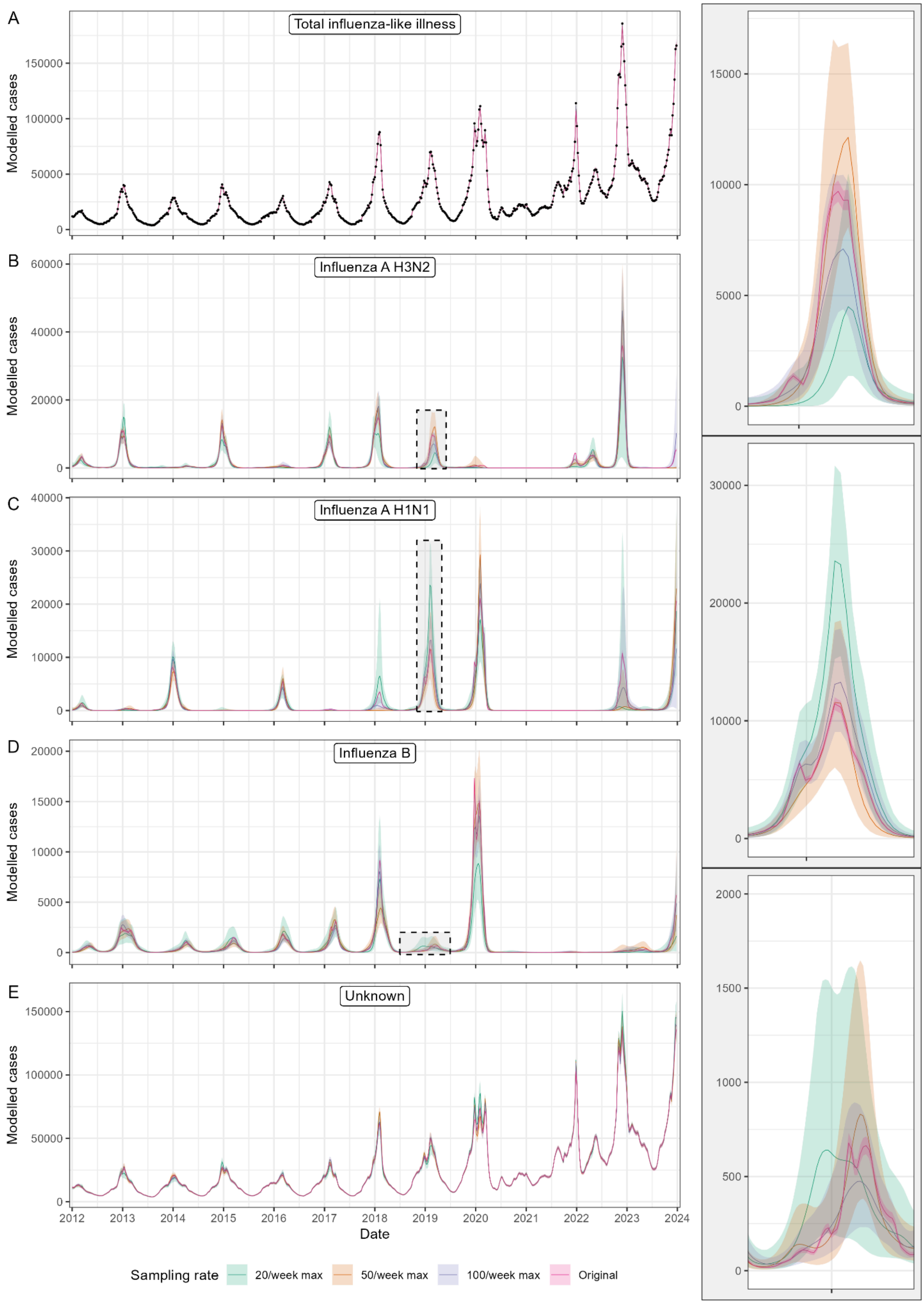


**SFig 11: Trends in influenza-like illness and influenza subtypes in the USA inferred at different testing rates.** (A) Modelled total number of weekly cases of influenza-like illness. (B-E) Modelled weekly number of cases attributable to influenza A H3N2 (B), influenza A

H1N1 (C), influenza B (D), and not attributable to influenza, ‘unknown’, (E). The three shaded boxes are shown magnified in the same order in the right-hand panels. For all panels modelled cases are shown for estimates made assuming cases of ILI were tested at the original sampling rate (Pink) or a maximum of 100 tests (Purple), 50 tests (Orange), or 20 tests (Green) a week. See SFig.2 for a visualisation of the testing data used at each sampling rate. All modelled estimates are shown with median (line) and central 50% (dark shaded region) and 95% (light shaded region) credible intervals.

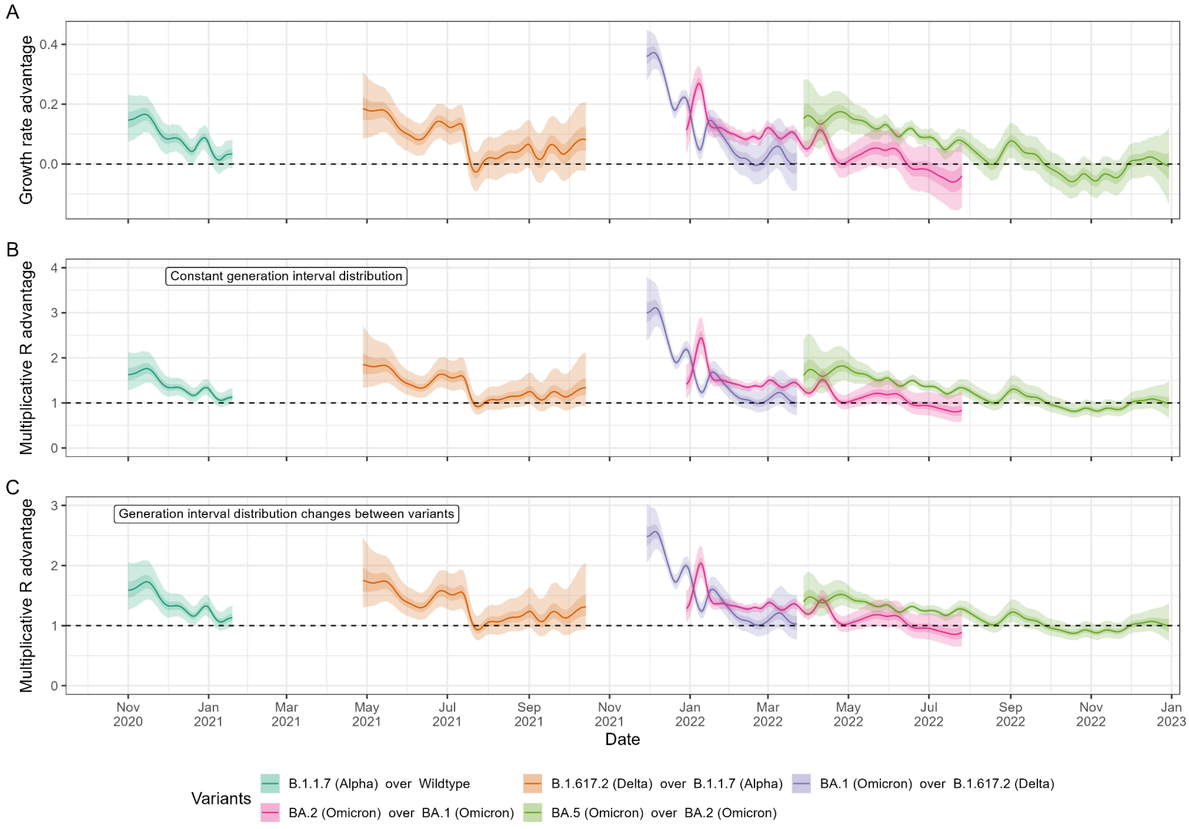


**SFig 12: SARS-CoV-2 variant advantages during periods of variant competition.** (A) Daily epidemic growth rate advantage between pairs of SARS-CoV-2 variants. (B) Daily multiplicative reproduction number advantage between variants assuming all variants had the same generation time distribution as wildtype lineages (SFig 13, STab 2). (C) Daily multiplicative reproduction number advantage between variants assuming all variants had different generation time distributions (SFig 13, STab 2). Estimates for the advantages of each pair of variants considered are only shown for the overlap of the periods of time after/before each variant's first/last detection in the UK. Wildtype lineages are only shown up to 19 January 2021 (when the rolling 7 day average for lineages classified as 'other' first dropped below one). All lineages categorised as 'other' prior to 19 January 2021 were wild type strains. All estimates are shown with median (line) and central 50% (dark shaded region) and 95% (light shaded region) credible intervals.

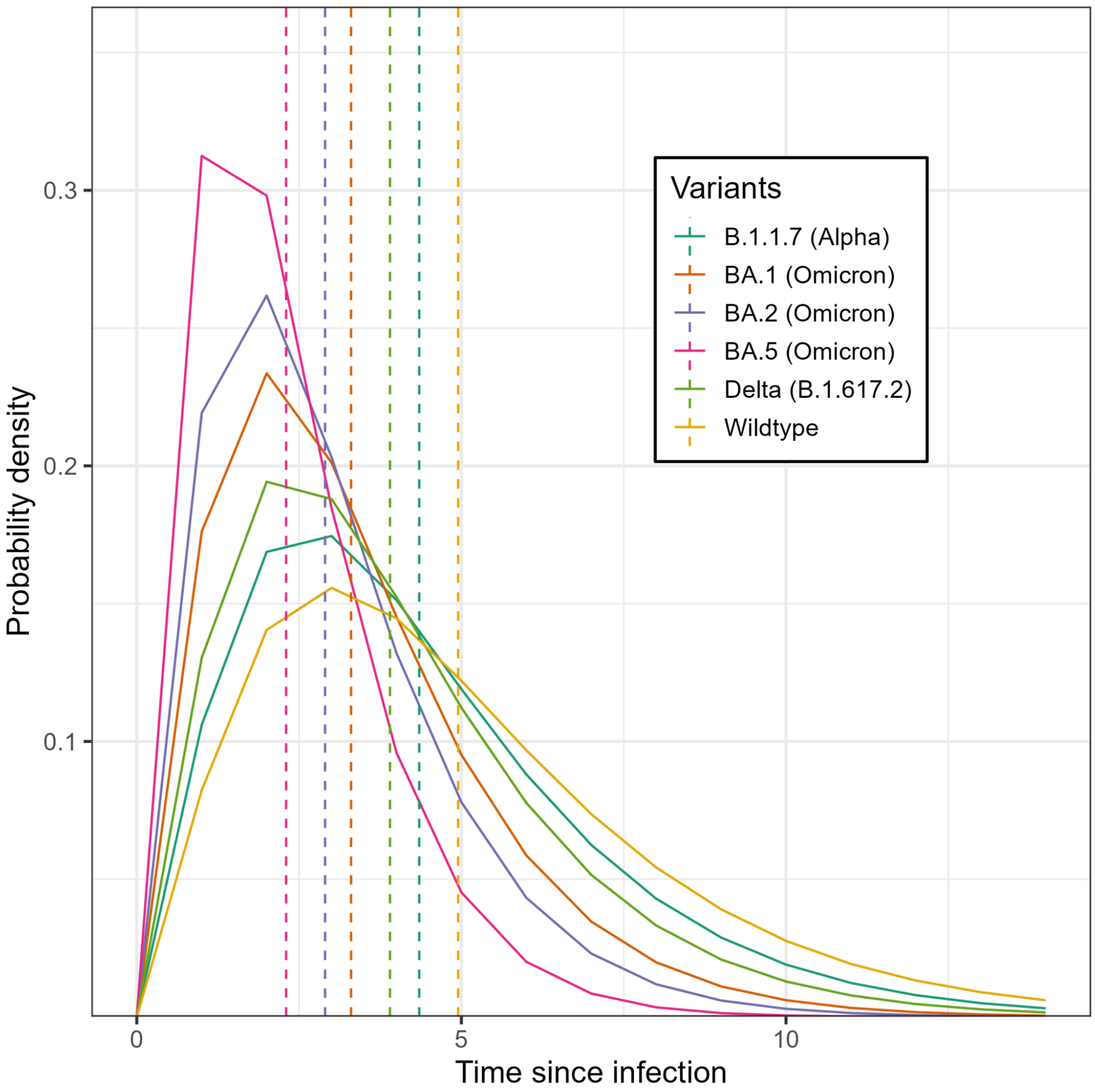


**SFig 13: Generation time distributions used for estimating each variant's reproduction number.** Generation time distributions (solid lines) used for estimating reproduction numbers, and the multiplicative reproduction number advantages in SFig.12B and SFig.12C. In SFig.12B all pathogens were assumed to have the same generation time distribution as wild type SARS-CoV-2, whereas in SFig.12C all variants had their own generation time distribution as shown. Also shown is the mean generation time (dashed vertical line).

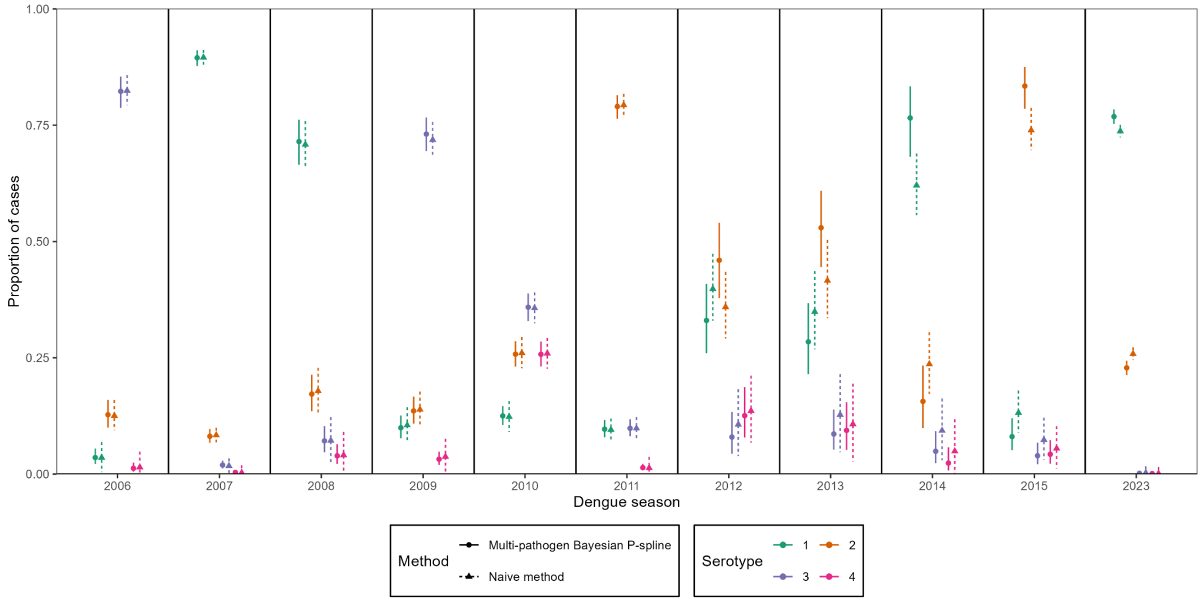


**SFig 14: The annual proportion of dengue cases caused by each dengue serotype in Taiwan (province of China).** The proportion of dengue cases caused by each dengue serotype for each season we considered. Each dengue season ran from 1 April to 31 March the following year. For example, the 2006 season ran from 1 April 2006 to 31 March 2007. We estimated the median (points) and 95% credible interval (error bars) in the proportion attributable to each dengue serotype using two methods: (1) using posterior modelled (multi-pathogen Bayesian P-spline) estimates of cases over time (solid circles, solid error bars); and (2) a naive method in which we calculate the proportion directly from the raw serotyping data for the entire season (i.e. fraction with exact binomial confidence intervals) which would not account for variable testing rates and case numbers.

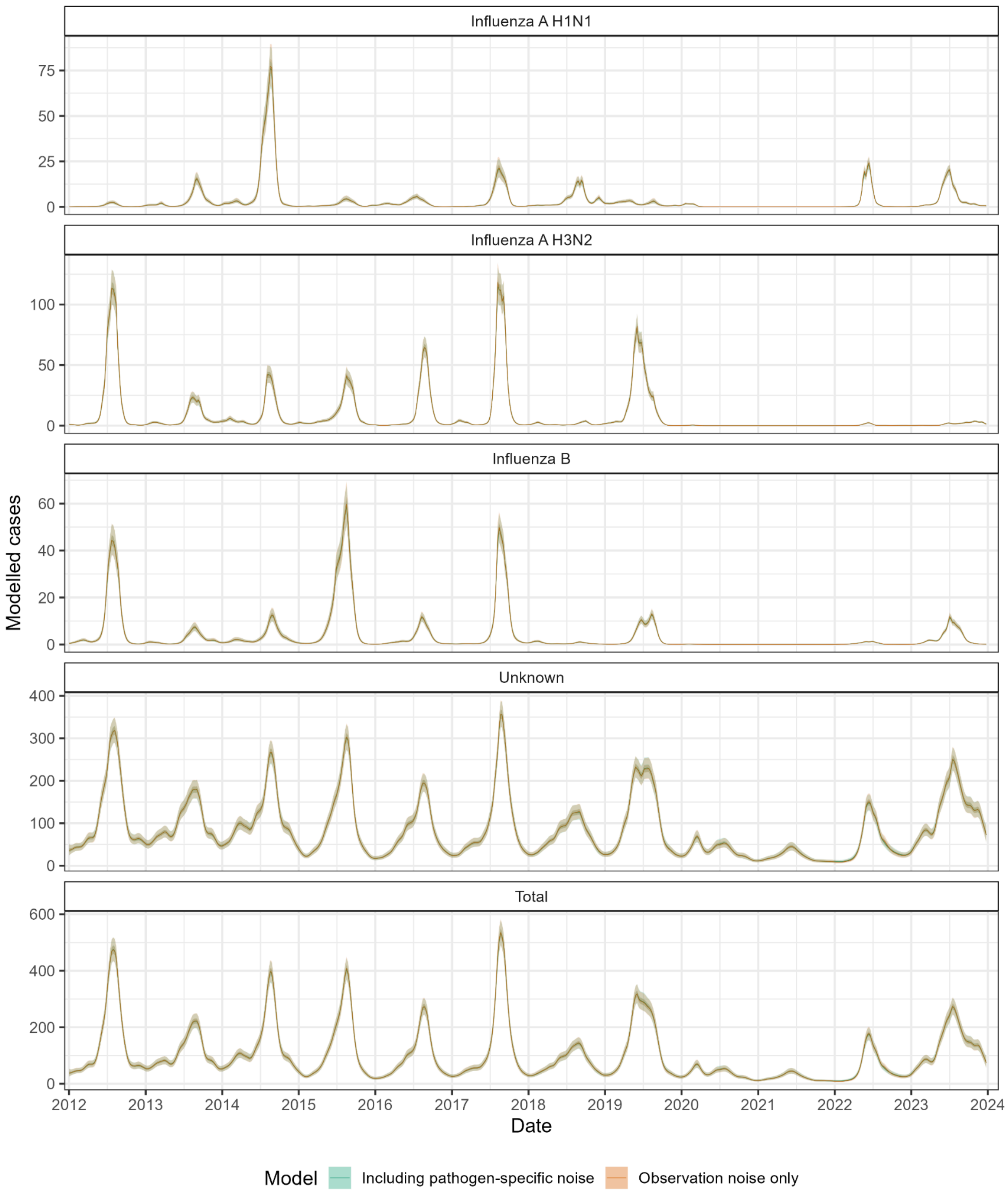


**SFig 15: Trends in influenza-like illness and influenza subtypes in Australia inferred when including additional noise terms in the model.** Modelled weekly number of cases attributable to influenza A H1N1 (first panel), influenza A H3N2 (second panel), influenza B (third panel), and not attributable to influenza, ‘unknown’, (fourth panel), and the modelled total number of weekly cases of influenza-like illness (bottom panel). All modelled cases are shown for estimates made using the original model (i.e. the one used for the main analyses) that just includes noise in the observation process (Orange), and the extended model that also includes a source of noise that is pathogen-specific (Green). All modelled estimates are

shown with median (line) and central 50% (dark shaded region) and 95% (light shaded region) credible intervals.

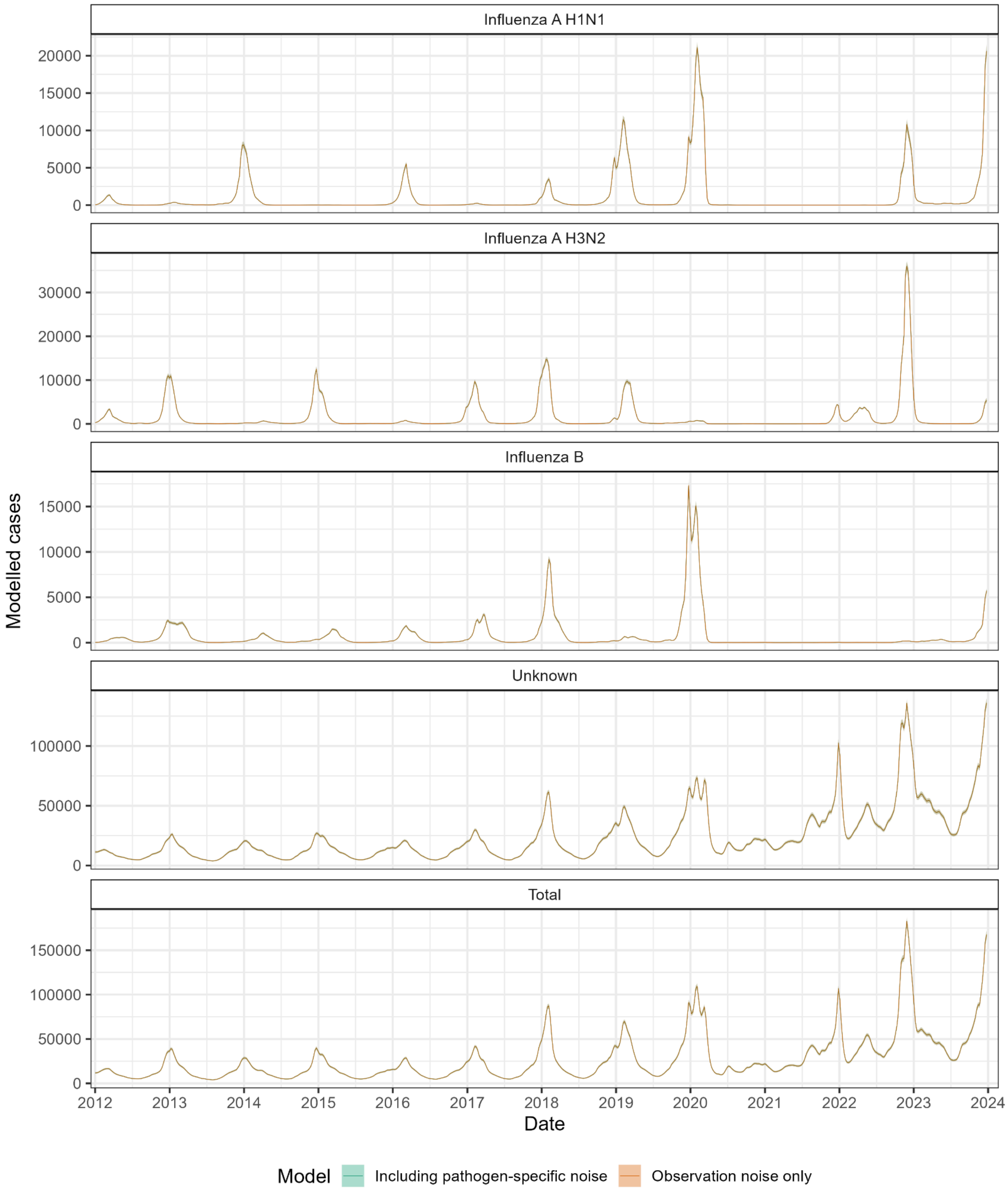


**SFig 16: Trends in influenza-like illness and influenza subtypes in the USA inferred when including additional noise terms in the model.** Modelled weekly number of cases attributable to influenza A H1N1 (first panel), influenza A H3N2 (second panel), influenza B (third panel), and not attributable to influenza, 'unknown', (fourth panel), and the modelled total number of weekly cases of influenza-like illness (bottom panel). All modelled cases are shown for estimates made using the original model (i.e. the one used for the main analyses) that just includes noise in the observation process (Orange), and the extended model that also includes a source of noise that is pathogen-specific (Green). All modelled estimates are

shown with median (line) and central 50% (dark shaded region) and 95% (light shaded region) credible intervals.

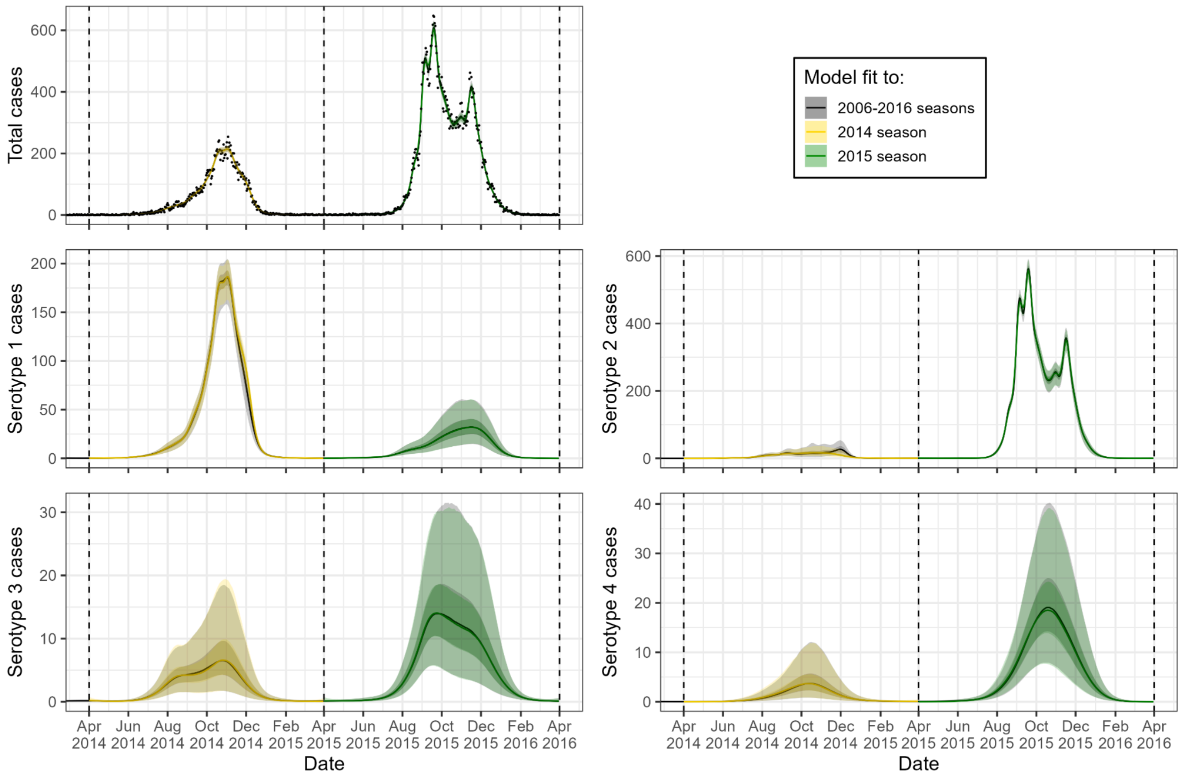


**SFig 17: Trends in dengue serotypes in Taiwan (province of China) inferred when fitting models to a single season with more informative priors.** (Top-left panel) Modelled total number of dengue cases (lines, and shaded regions) and daily number of dengue cases (black dots). Modelled daily number of dengue cases attributable to each dengue serotype, and modelled total number of dengue cases for the 2014 and 2015 dengue seasons. The daily number of dengue cases (points) are also shown in the top-left panel. Modelled estimates are shown from models fitting to the 2014 season only (Yellow), and 2015 season only (Green) using prior distributions informed from a model fit to the 2006-2013 seasons (not plotted). Modelled estimates from a model fit to the 2006-2015 seasons without an informative prior (Grey) are also shown for comparison. Note that there is almost complete overlap in the posterior estimates. The dashed lines show the start date of each dengue season (1 April). All modelled estimates are shown with median (line) and central 50% (dark shaded region) and 95% (light shaded region) credible intervals.

**Supplementary tables**

**STab 1: Description of model assumptions for all analyses performed in this paper.**

**STab 2: Generation time distributions assumed when estimating multiplicative reproduction number advantages for SARS-CoV-2 variants.**
